# Interpretable and Predictive Deep Modeling of the SARS-CoV-2 Spike Protein Sequence

**DOI:** 10.1101/2021.12.26.21268414

**Authors:** Bahrad A. Sokhansanj, Zhengqiao Zhao, Gail L. Rosen

## Abstract

As the COVID-19 pandemic continues, the SARS-CoV-2 virus continues to rapidly mutate and change in ways that impact virulence, transmissibility, and immune evasion. Genome sequencing is a critical tool, as other biological techniques can be more costly, time-consuming, and difficult. However, the rapid and complex evolution of SARS-CoV-2 challenges conventional sequence analysis methods like phylogenetic analysis. The virus picks up and loses mutations independently in multiple subclades, often in novel or unexpected combinations, and, as for the newly emerged Omicron variant, sometimes with long explained branches. We propose interpretable deep sequence models trained by machine learning to complement conventional methods. We apply Transformer-based neural network models developed for natural language processing to analyze protein sequences. We add network layers to generate sample embeddings and sequence-wide attention to interpret models and visualize multiscale patterns. We demonstrate and validate our framework by modeling SARS-CoV-2 and coronavirus taxonomy. We then develop an interpretable predictive model of disease severity that integrates SARS-CoV-2 spike protein sequence and patient demographic variables, using publicly available data from the GISAID database. We also apply our model to Omicron. Based on knowledge prior to the availability of empirical data for Omicron, we predict: 1) reduced neutralization antibody activity (15-50 fold) greater than any previously characterized variant, varying between Omicron sublineages, and 2) reduced risk of severe disease (by 35-40%) relative to Delta. Both predictions are in accord with recent epidemiological and experimental data.

## INTRODUCTION

The emergence of the COVID-19 pandemic has coincided with the widespread availability of lower cost, rapid whole genome sequencing. While sequencing is still not as globally equitable or ubiquitous as it should be, an unprecedented amount of sequence data has been generated for SARS-CoV-2, the novel coronavirus that causes COVID-19. As of October 2021, when the data for most of the studies in this paper was collected, nearly 4 million sequences were available to researchers from the GISAID website (http://www.gisaid.org). [1] As of December 22, 2021, that number has grown to nearly 6.4 million sequences. The key challenge now is how to translate abundant biological sequence data to as much biological *information* as possible. This goal is particularly urgent as the virus continues to rapidly mutate and change, often in fundamental ways that affect disease severity and transmission—and experimental tools other than sequencing are costly, time-consuming, and often technically difficult to perform.

Conventional ways of interpreting large scale sequencing data rely on phylogenetic methods or sequence alignment and comparison to 1) group viral sequence variants into strains or lineages whose properties can be measured collectively, and 2) identify specific mutations with potential impact on virulence, transmission or immune escape. The quantity, diversity, and pace of changes in the SARS-CoV-2 genome as the pandemic has expanded—driven by its novelty to the human immune system and sheer number of infections worldwide—has strained conventional approaches to their limits. [2] While phylogenetic methods have been powerful tools to trace the transmission and origins of SARS-CoV-2, as the pandemic has expanded, the virus has picked up and lost mutations, often in novel or unexpected combinations. [3] When genetic lineages or subclades have reached a high level of fitness and spread around the globe, such as Alpha and Delta, complex sublineages have emerged which may have different immune evasion and virulence properties, sometimes sharing more in common with other genetic lineages than their parents. And, because of the quantity and diversity of sequencing that has been done during the pandemic, we can see the complex genetic evolution of the virus in real time. Using alignment to identify single site mutations has also proven limited. Changes to SARS-CoV-2 properties can implicate complex combinations of multiple mutations that emerge simultaneously—and then sometimes even revert as the virus continues to evolve. [4]

We propose a framework of deep neural network sequence models as a complement to conventional comparative genomics techniques. In particular, we apply Transformer-based neural network models developed for the natural language processing field [5] to analyze protein sequences, by representing amino acids similarly to how language processing models represent words in a sentence. We further include layers in the neural network to generate spike protein embeddings and sequence-wide attention mapping in addition to making predictions. The embeddings and attention provide information that can be used to interpret how the model uses sequence information to make predictions, and to find deep patterns and connections within sequence that are relevant to the function of viral proteins.

In this paper, we demonstrate our interpretable deep modeling approach of the SARS-CoV-2 spike protein, which is responsible for binding to host cell receptors and mediating and cell entry, and which is a major target for host immune response. [6] While we focus on SARS-CoV-2 and the spike protein in this paper, the model architecture we develop can be flexibly adapted to other biological sequence problems. Indeed, we show that within the SARS-CoV-2 spike protein context, where the sequence length remains approximately constant, we can train the model on diverse classification and regression tasks without changing the model architecture or going through costly hyperparameter tuning exercises. These tasks include: (i) regression of sequence data to the date of a sequence’s first emergence, as a way to qualitatively validate the significance of the embedding and attention-based interpretations generated by our model; (ii) predicting disease severity of sequences by using patient metadata available for a subset of sequences in the GISAID databases and using attention to identify mutations that may be related to heightened risks of severe disease; and (iii) demonstrating the flexibility of our approach by applying it to genus-level classification of coronavirus spike proteins.

While this manuscript was being finalized, a new SARS-CoV-2 lineage, B.1.529, was identified as a variant of concern by WHO and named Omicron. First reported on November 25, 2021 from sequences in Southern Africa, the Omicron variant has exhibited rapid growth around the world. [7] Because we did not consider any sequence data in GISAID after October 2021, the other studies in this paper entirely excluded Omicron sequences. Omicron also contains numerous spike protein mutations. [8], [9] Given that many of these mutations are in the spike protein’s receptor binding domain, it raises important questions about Omicron’s ability to evade the immune response in previously infected and vaccinated patients, as well as infectivity and virulence. Consequently, Omicron is a critical challenge for our modeling approach. The Omicron variant has different sequence properties from other variants found before its emergence, so it differs significantly from the data used to train the neural network model. We demonstrate that the model can be used to make computational predictions before any empirical data emerges, specifically by predicting Omicron’s immune evasion and relative risk of disease severity as compared to Delta and Alpha. This proof of concept shows the potential for a predictive computational model that can provide a starting point for epidemiological guidance before health observations and laboratory data are collected.

## 2. BACKGROUND & RELATED WORK

### Evolution of Coronavirus lineages and the SARS-CoV-2 spike protein

We focus on the spike protein because of its crucial role in COVID-19 infection and the immune response. [6], [10], [11] As a practical matter, moreover, the spike protein is the key target for COVID-19 vaccines and antibody therapies. [12]–[14] Briefly, the SARS-CoV-2 spike protein (S) is a glycoprotein that is responsible for binding to the host cell receptor, angiotensin-converting enzyme 2 (ACE2) and then gaining entry to the cell. [6] The spike protein is responsible for cell membrane entry in all coronavirus species. Generally, the spike proteins of coronaviruses have two subunits, S1, which is responsible for binding to the receptor, and S2, for membrane fusion. [15] The sequence of the SARS-CoV-2 spike protein is similar to that of SARS-CoV-1 (76% identity), and more than 90% identity with bat and pangolin coronavirus species, which may suggest its origin in those animals. [6] Notably, even when the spike protein binds the same receptor (e.g., ACE2 in the case of SARS-CoV-1 and SARS-CoV-2) there are differences in antigenicity and epitopes. [16] Collectively, there are four genuses of coronavirus, *Alphacoronvirus, Betacoronavirus, Gammacoronavirus*, and *Deltacoronavirus*. The coronavirus species known to infect humans are all betacoronaviruses, but lie in different subgenuses: SARS-CoV-1 and SARS-CoV-2; MERS-CoV; and HCoV-229E, HCoV-OC43, and HKU-1 respectively; the last pair of which generally cause mild disease. [17] Coronavirus spike proteins bind a variety of different receptors. [18] Indeed, in addition to ACE2, SARS-CoV-1 also binds dipeptidyl peptidase 4 (DPP4); however, SARS-CoV-2’s spike protein only weakly binds DPP4 and there is no evidence that it uses it as a host cell receptor. [19] Other differences in spike protein sequence can influence how the coronavirus interacts with the host cell as well. For example, unlike its closer genetic relatives, SARS-CoV-2 spike has a furin cleavage site, which is used to prime for cell entry. [20] In sum, relatively small differences in spike protein sequence can have substantial effects on the protein structure and function.

As the COVID-19 pandemic has continued, the SARS-CoV-2 spike protein itself has accumulated mutations, and variant protein sequences have emerged. As it is a RNA virus, SARS-CoV-2’s genome is prone to mutate, albeit at a rate is mitigated by its large genome size and the proofreading function of the exoribonuclease that it encodes. [21] The most frequent mutations observed in coronaviruses more generally are substitutions, although insertions and deletions are observed as well. [22] In some cases, insertions from other viral genomes may occur, and, in fact, it appears as though the SARS-CoV-2 genome includes an insertion from human RNA. [23] In other human coronaviruses, estimated mutation rate is around 3 × 10^−4^ substitutions per site per year. [24], [25] Early on in the pandemic, spike protein sequence variants began to be observed, such as a substitution D614G, which rapidly became dominant as the pandemic began to spread within Europe. [26], [27] Since then, the amount of mutation observed during the COVID-19 pandemic has been more substantial, perhaps because it is a novel virus exposed to hosts around the world. [28] We have seen that SARS-CoV-2 variants can have critical differences in the level of virulence, pathogenicity, immune evasion (with implications for therapeutic antibodies and vaccine efficacy), and transmissibility. [2]

Because of its critical role in antigenicity and cell entry, changes in the spike protein sequence can be particularly impactful. And the SARS-CoV-2 spike protein will continue to change. Even before the emergence of the novel coronavirus, it was shown that in another human coronavirus, HCoV-OC43, the spike gene had a higher rate of substitution and positive selection sites than other genes. [29] This suggests that genetic drift in the spike gene plays a role in adaptive evolution of HCoV-OC43 and likely also other coronaviruses, given that the spike protein is the major antigen recognized by the immune response. Indeed, one paper published in July 2021 estimated that, to that point, SARS-CoV-2 had only “explored” 31% of the potential space for spike protein variation, based on comparisons with related sarbecoviruses. [30]

Initial work on the impact of spike protein variants focused on the D614G substitution in the spike protein, which was found in an animal model to have an increase in fitness and transmissibility. [31] However, as other studies accumulated, it proved difficult to show a clear impact on virulence or transmissibility, although clinical samples with the variant were found to have higher virus titers. [32] The wide spread of the Alpha (B.1.1.7) variant starting in December 2020 did suggest that it had an increase in transmissibility over wild type and previous variants [33], although it is unclear that it resulted in more severe disease. [34]–[37] The later Delta (B.1.617.2) variant, however, has had a clear-cut increase in transmissibility supported by both epidemiological estimates [38] and laboratory studies that show increased fitness over previous variants, including enhanced viral replication due to modification in the furin cleavage site of the spike protein. [39], [40] Epidemiology and cell/animal-model work has been complemented by computational and in vitro work mapping the impact of individual mutations on spike protein structure, fitness, and antibody evasion. [41], [42] One area in which intensive study of the effect of variants has been undertaken is in studies of potential impact on vaccine efficacy. Particular variants, such as the Beta (B.1.351) variant once dominant in South Africa, have consistently shown substantial decreases in neutralizing antibody response in sera from individuals vaccinated with diverse adenoviral vector, mRNA, and recombinant protein subunit vaccines. [43]–[48] This has been paralleled by reduced efficacy in clinical trials and studies among patients with sequenced variants and in South Africa. [49], [50] Moderately reduced neutralizing antibody response was also found for the Delta (B.1.617.2) variant, which became dominant worldwide through 2021. However, the impact of vaccine efficacy turned out to be proportionately greater, perhaps due to Delta’s increased replication proficiency. [40], [51] These differences in vaccine efficacy are consistent with experiments which show that the principal SARS-CoV-2 variants observed so far, including Alpha, Beta, and Gamma, are antigenically distinct; in particular, that convalescent sera from patients infected with one of these variants shows the smallest reduction in neutralizing antibody to the wild type of the virus and greater reductions to other variants. [52]

Fortunately, widespread genetic sequencing has allowed for an unprecedented view into changes in the SARS-CoV-2 genome over time and in different regions. [53], [54] Over five million sequences are now available in the GISAID (Global Initiative on Sharing All Influenza Data) database (http://www.gisaid.org), which has encouraged data sharing by trading restrictions on republishing raw sequence information with access to that information for analysis. [55] So much data has been generated and made available that it has spurred the development of computational tools for high frequency tracking [56] and even daily updates.

[57] The first prominent tool for tracking SARS-CoV-2 genomic variation was the NextStrain project, which was originally developed as a general tool for viruses and adapted to offer clade definitions for SARS-CoV-2 based on phylogenetic analysis. [58] Subsequently, the Pango lineage (Pangolin) definition of lineages was developed, with an emphasis on highlighting potentially significant lineages that would be grouped together, supported by a machine learning approach (using random forests) to classify sequences to lineages. [59] Particularly significant Pango lineages have been identified by the World Health Organization (WHO) as variants of concern (VOC) and given Greek letter designations [60], such as Alpha (B.1.1.7), Beta (B.1.351), Delta (B.1.167.2), and, recently, Omicron (B.1.1.529).

Being able to leverage the sheer volume of available data has proven to be a challenge. Some have questioned whether the current methods of classifying variants is sustainable as so many have emerged. [2] One drawback for tree-based approaches to classification is that particular variants may occur independently in different lineages, as has been observed, for example, in the N501Y substitution which has arisen in multiple clades (without any evidence for recombination). [61] Another challenge for a phylogenetic approach is that multiple variants may occur within an individual infection. [62] Some immunocompromised individuals have chronic infections that can last six months to a year [63] In some cases, those patients may be treated with convalescent plasma or antibodies which may be selecting for evasive mutations that are being observed. [64] During the long-term infection, a spike protein can emerge with multiple variations, which phylogenetic analysis identifies as “long branch” divergence from the phylogenetic tree. Indeed, the Omicron variant has such a long branch divergence, which has led to suggestions that it may have emerged in an immunocompromised host, or even potentially reverse zoonosis given that COVID-19 can infect animals as well. [9]

The challenges that SARS-CoV-2 evolution pose to conventional computational methods have motivated the development of alternatives based on bar-coding [65], co-mutational modules [66], and k-mer abundance [67]. However, even these methods may be unsustainable as they are based on historical information, even as phylogeny-based classifications become more chaotic. [68] Further complicating the problem is that combinations of variants can have nonlinear effects, either synergistic, antagonistic, or neither. In one study of antibody responses in convalescent plasma from recovered patients and sera from vaccinated but uninfected individuals, it was found that a simultaneous deletion at sites 69-70 (ΔH69/V70) as well as substitution E484K had enhanced decrease in antibody neutralization. Conversely, L452R and P681R resulted in a decreased neutralization less than the effect of both individually. Another pair, E484K and N501Y, resulted in a neutralization decrease that was the sum of each. [69]

### Deep Learning’s potential and pitfalls in predicting outcome and evolution of SARS-CoV-2

Within the more than five million sequences uploaded to GISAID, a small fraction (nearly 220,000 of over 5,500,000 sequences as of November 27, 2021) have some metadata about the clinical status of patients. Such metadata can include whether the sample comes from someone who is alive or dead, whether they were in the ICU or had mild or severe disease, etc. Generally, only one piece of information (such as “alive” or “dead,” or “mild” or “severe”) is conveyed through the metadata annotation. The GISAID dataset and focused patient studies (which are often the source of the data available on GISAID) have enabled researchers to study the potential link between sequence variation and virulence. These studies have included analysis of single nucleotide polymorphisms (SNP) and genome wide association studies [70], [71], literature meta-analysis [72], genome wide association studies [73], and the incidence and prevalence of mutations in GISAID entries with clinical metadata [74], [75]. The results of these studies found some correlation of symptomatic or severe disease with spike variants such as D614G, however, none of these have been verified elsewhere. However, another analysis of country-based case fatality rates (CFR) found no relationship between mortality and clade, at least through March 2021. [75] To the extent that patient status in GISAID metadata does show variation, other studies using logistic regression and conventional statistical techniques found it to largely reflect worse outcomes for older males, consistent with commonly understood clinical experience, while it was difficult to discern variation at the clade or sequence levels. [76], [77] Other more complex analysis has included the use of deep neural networks, in particular, a combination of convolutional neural network (CNN) and recurrent neural network (RNN), which attempted to predict what mutations would increase virulence severity based on countrywide death statistics. [78] Another approach that has been developed and recently validated is the use of Bayesian multinomial logistic regression to infer growth rate from individual mutations; from that, the authors posited that the mutations correlated with high growth rate also conferred fitness and transmissibility benefits. [79]

The complexity and volume of sequencing data have motivated the application of deep learning methods based on applying natural language processing (NLP) methods to DNA and protein sequence data. These works exploit the analogy between the semantic structure of language and the bases and amino acids that make up nucleotide and protein sequences. For example, one group has used concepts from semantic processing, e.g., the frequency of correlated words, to identify potential mutagenic sites in viruses including SARS-CoV-2. [80] A related work has used deep learning methods to simulate coronavirus spike protein variation to predict potential zoonotic sequences. [81] Another group has applied “ProtBERT” protein sequence embeddings obtained through self-supervised learning applying the BERT (Bidirectional Encoder Representations from Transformers) method from NLP to SARS-CoV-2 in combination with k-means clustering to identify groups of correlated mutations. [82] Another work applied Long-Short Term Memory RNNs (LSTM) to classify sequences to lineages. [83]

While deep learning methods can identify complex features within data that allow classification, that strength comes with a major weakness: understanding what the deep model focused on in learning the classification and explaining its predictions. Accordingly, interpretable machine learning has emerged as a significant area of research. [84], [85] Interpretable modeling is a critical need in biological sequence analysis. [86], [87] Interpretable modeling allows us the biomedical community to justify high stakes clinical and research decisions based on machine learning predictions. Interpretable models also allow researchers to leverage data to obtain as much understanding as possible, given that other biological techniques are often costly, slow, and hard to do reliably. Proposed methods for interpretable machine learning include examining the neural network structure, through relevance propagation, propagating activation differences through the network, and sensitivity analysis methods. [88], [89] Integrated gradients have been used in bioinformatics for developing models for RNA splicing. [90] For CNN-based models, a saliency map method has been proposed for identifying what part of a sample (image, or text) was identified as an important feature by the model. [91]

In view of the analogy between biological sequence and written language, we can also look to methods developed for NLP. In particular, attention mechanisms developed by [92] and [93] have been used to highlight the important features that deep learning methods used to generate text classification models. [94]–[96] Deep learning models combining CNN with attention have been used to identify sequence motifs for functional genomics, e.g., transcription factor binding site detection. [97], [98] Another work generated predictive models of adverse drug reactions based on chemical structures by combining attention with a CNN for each chemical property and structural feature in the model. [99] We have shown that attention in combination with a LSTM-based sequence model can be used to obtain insight into taxonomic and phenotypic classification of 16s ribosomal RNA sequences of bacteria [100], as well as gene ontology classifications of protein sequences [101]. Recently, transformer-based architectures, which are built on multiple attention modules, have emerged as being important in NLP. The attention within transformers has been proposed as a source of explainability. For example one work demonstrated how different attention heads attended to different aspects of a learning task to identify nucleotide motifs for promoter sequences. [102] However, attention cannot be inherently drawn out of transformer-based architectures, and it has been suggested that further processing steps may be required to connect attention to specific linguistic features. [103] It should further be noted that attention is not necessarily tied to explanation, in the sense of explaining why a prediction took place, but it can highlight features that the model paid attention to at the attention layer making a prediction, although those may not have been the important features with respect to the ultimate classification problem. [104]

## METHODS

### Data Collection and Pre-Processing

As described further in the Results & Discussion section, the studies shown in this paper employ three different kinds of coronavirus spike protein sequences: SARS-CoV-2 spike protein sequences submitted to the GISAID database through the course of the pandemic, protein sequences which were provided to GISAID with patient metadata information specifically, and protein sequences from coronavirus of all genera. The procedures for collecting and processing sequence data are outlined for each type below in turn.

#### SARS-CoV-2 Spike Protein Sequences

We downloaded a FASTA file of protein sequences from the GISAID database, http://www.gisaid.org. (See Supplemental File and for a list of acknowledgments to contributing laboratories.) The spike protein sequences are preprocessed by GISAID by multiple sequencing alignment, identifying ORFs, and translating nucleotide sequences to obtain protein FASTA files. We then parse the FASTA file to obtain a file with only the Spike protein sequences. (GISAID also offers a FASTA file of only Spike protein sequences; however, that was not used for this paper.) Unless indicated otherwise, the protein sequences we use for our analyses shown are those submitted and preprocessed by GISAID as of October 16, 2021. The metadata annotation file from GISAID including sequences collected and processed as of October 1, 2021 was also downloaded from the GISAID website. This metadata file included the collection date, annotated Pangolin lineage, Nextstrain-identified clade, and geographical information.

In this paper, we at times refer to “raw” and “aligned” sequences. “Raw” sequences are exactly those that were contained in the FASTA file downloaded from GISAID. “Aligned” sequences are generated using the local pairwise Striped Smith-Waterman (SSW) method [105], [106], with BLOSUM62, in the scikit-bio package in Python 3.8. [106] Following alignment, we obtain a protein sequence in which all insertions and deletions are positioned with respect to the consensus Spike reference sequence (Wuhan-Hu-1 isolate) which was obtained by multiple sequence alignment of early genome sequences. [107] Variant single site polymorphisms can then be identified in accord with convention. First, the sites in the aligned data sequence corresponding to a gap in the aligned reference sequence were shifted such that any insertion was relative to the reference. Aligned sequences are then padded at the beginning and end of the aligned region (if it was less than the 1273 residue length of the reference sequence) with a mask (“*”) to formulate a 1273-residue long sequence. While we generally did not do so for the studies presented in this paper, it is also possible to “filter” aligned sequence data by removing any sequences containing any “*” (mask) or “X” (ambiguous amino acid) characters. We provide the source code for the alignment and processing of aligned sequences at https://github.com/bahrad/Covid.

#### GISAID sequences containing patient (clinical) status metadata

Our analysis considers sequences for which patient metadata were available on GISAID as of October 4, 2021. These sequences are downloaded in batches from the GISAID website and combined into a single CSV file of patient data. As of that date, there were 155,545 records indicated as having “patient metadata” available from GISAID. Patient metadata on GISAID consists of a single field, which includes some text provided by the submitter of the sequence. We assign raw text in the metadata field of each record to one of the following categories, according to the scheme in Supplemental Table 1: Alive, Asymptomatic, Dead, Hospitalized, Mild, Moderate, Released, Screening, Severe, Symptomatic, and Unknown (which includes any other categories identified in Supplementarl Table 1, such as “Vaccinated”). These categories follow the commonly used case classification such as those defined by the United States National Institutes for Health (NIH) COVID-19 guidelines. [108] For example, metadata indicating ICU admission is categorized as “Severe.” We remove all data that corresponding to categories Alive, Symptomatic, and Unknown, as they cannot be further classified as “Mild” or “Severe.” For example, a “Symptomatic” or “Alive” patient may have severe symptoms or have been hospitalized. We assign the following categories to the classification of “Mild,” or the numerical value of 0: Asymptomatic, Mild, Moderate, and Screening. We assume that the last category was not identified as a result of reporting symptoms or at least seeking hospitalization due to Covid symptoms as opposed to incidental to another condition). We assign the following categories to “Severe,” or numerical value of 1: Dead, Severe, Hospitalized, and Released. The last category indicates prior hospitalization. Finally, we also remove all samples which are not explicitly identified as being from a human patient, as identified in GISAID’s host metadata field.

The resulting metadata records are then merged with the aligned sequence data set obtained from GISAID, preprocessed as described above. The training data set is made up of samples that were submitted and processed in time to be available as of September 12, 2021, as well as having patient metadata available as of that date. The samples used in the training data set are those with sequences that were found at least five times overall in the sequence data prior to September 12, 2021. The remaining samples that were found in the October 1, 2021 dataset from GISAID that had patient metadata available as of October 4, 2021 make up an independent validation set. This data set includes that were collected after October 1 and contain sequences that were not found at least five times prior to September 12. As a result of the aforementioned procedure, there is no exact sequence overlap between training and test samples.

As further described in the Results & Discussion, it is necessary to account for potential confounders of age, gender, and sample collection date relative to the onset of the pandemic. We therefore process the age metadata to remove any unknowns or text entries which cannot be identified as an age. We also remove any samples with a clearly incorrect date, i.e., predating the onset of the pandemic in late 2019, and any samples with missing or nonreadable date metadata. We also convert the dates to days since December 1, 2019, which predates the first sequenced samples from COVID-19 patients. For gender, we convert male and female metadata to integers 0 and 1 respectively, and we remove all samples with unknown or ambiguous gender metadata. Finally, each sample is associated with a vector of three integers, corresponding to age, gender, and sample date, which is input to our neural network model at the *N*_*H*_-dimensional embedding layer as shown in Fig. 1. We do not remove samples with incomplete dates, instead we assign them to the latest possible date.

**Figure 1.**
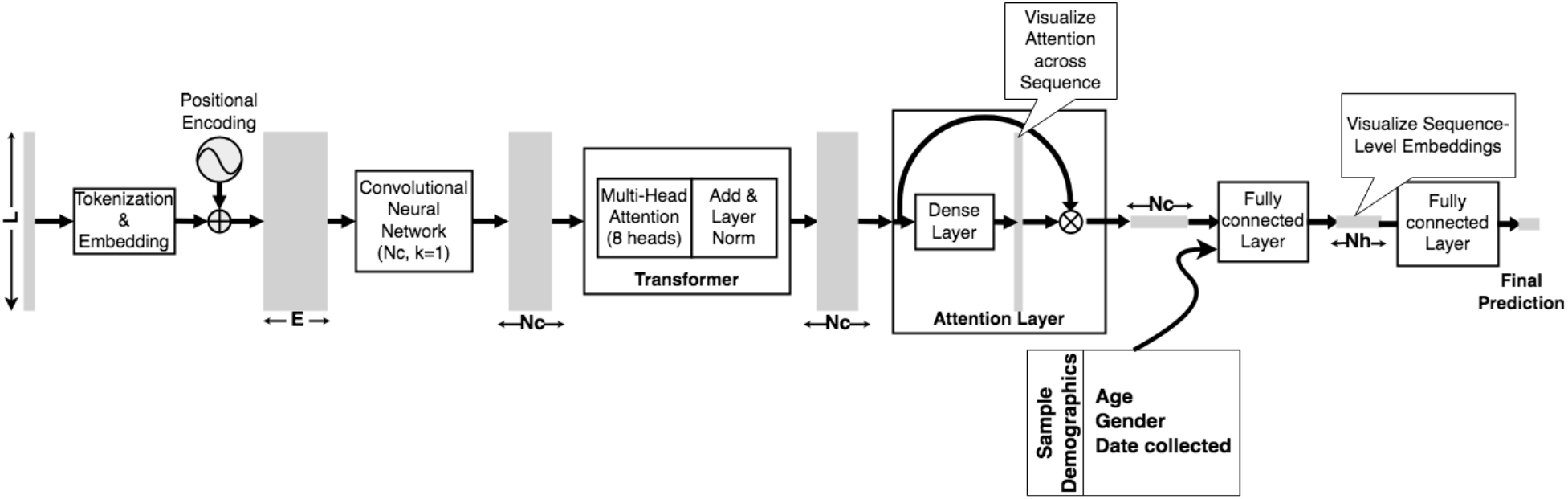
Neural network architecture used in this paper. The sequence of length *L* is given a token and position embedding, which is then compressed using a CNN layer with *Nc* filters prior to going to a multi-head attention Transformer encoder. The encoder output is then processed by a self-attention layer, which allows for visualization of attention along the *L*-length sequence. The output of the attention layer is then optionally concatenated with additional variables encoded in nonzero integers, i.e., demographic variables as shown here, and input to a densely connected layer with *N*_*H*_ nodes. A resulting “embedding” of the sequence may be visualized at this stage, which will be a vector of (usually) *N*_*H*_ << *L* dimensions. Finally, the prediction is provided by either a clipped linear or sigmoid node (for regression), a sigmoid node (for binary classification), or a softmax (for multiclass classification).

#### Taxonomic classification of multi-species spike protein sequences

For the study of taxonomic classification of coronaviruses across all genera (*Alpha-, Beta-, Gamma-*, and *Deltacoronavirus*), we use spike protein sequences downloaded from the NCBI Virus website, https://www.ncbi.nlm.nih.gov/labs/virus/vssi/. The spike protein sequences used in this study were downloaded on November 21 and 27, 2021 using search terms such as “spike,” “S1 protein”, “S2 protein,” and “S protein,” in order to download as many spike protein sequences as possible. Sequences that are shorter than 1000 amino acids in length or which did not have coronavirus as an identified species are then removed. Additionally, in order to make the SARS-CoV-2 classification task less trivial, we removed the sequences that are known to have the closest matches to SARS-CoV-2 spike protein, which are SARS-like sequences, such as the WIV-1 bat coronavirus, any recombinant viruses that were generated to research SARS-CoV-2, and sequences identified being from “SARS-like” viruses in the database. In sum, 18,026 sequences are downloaded from NCBI, and, after removals, the resulting data set consists of 7,544 sequences.

To evaluate the taxonomic classifier, we use three data sets: 1) a random sample of 100,000 of the over 3 million raw spike sequences obtained from GISAID as described above, 2) all of the raw spike sequences with a length shorter than 1000 amino acids, and 3) all raw sequences with more than 250 “X” residue sites, where a unique amino acid could not be identified.

#### Predicting the Omicron variant’s capacity to escape immune/vaccine response

For Omicron, we develop a training data set based on the reduction in neutralizing antibody titers to variant viruses in sera from vaccinated patients as presented in the literature, as shown in Table 1. [109]–[113] As described further in the Results & Discussion section, we needed to integrate data from multiple studies, which may involve different experimental conditions such as whether they used native virus or pseudovirus (e.g., HIV-1 engineered to express spike proteins with a particular variant sequence) and what kinds of vaccinated sera were compared. Where possible, we used data based on reductions of neutralizing antibodies for sera from patients vaccinated with the Pfizer/BioNTech BNT162b2. We use the aligned data set described above to source the training data set. We include sequences that are annotated with the lineages for which there is information in Table 1 and assign them as a label the relative neutralizing antibody activity from Table 1. We remove all sequences that occur fewer than 3 times in the database overall, and then compose the training data set based on up to the 300 most frequently occurring sequences for each of the labels. The result is a training data set of 2,245 samples. We then balance classes to account for those labels which have fewer than 300 representatives using `class_weight.compute_class_weight` in scikit-learn (sklearn) version 1.01, which obtains the weights of samples in a class by dividing the average number of samples in each of all classes by the number of samples in that class. [114]

**Table 1.** Training data used for model prediction of Omicron immune evasion. Sequences with the lineages indicated in the table are assigned the label of the reciprocal of the corresponding fold decrease in antibody neutralization. (See citations in accompanying text.) Where there is a WHO-identified variant of concern, the variant identity is provided. B.1 is the first variant to become widespread, arising from the wild type emerging from Asia and spreading throughout Europe and North America. It is largely the same as the wild type, with the D614G modification as most important. Delta-like lineages B.1.617.1 and B.1.617.2+ are included as well; B.1.617.2+ is sometimes called “Delta+” (although other lineages have been called that as well). [115] In this study, B.1.617.2+ are sequences annotated as B.1.617.2 but which also include the E484K mutation.

| Lineage (Variant) | –Fold decrease in Ab neutralization |
| --- | --- |
| B.1 | 1 |
| B.1.1.7 (Alpha) | 2.3 |
| B.1.351 (Beta) | 8.8 |
| P.1 (Gamma) | 2.9 |
| B.1.617.2 (Delta) | 3.8 |
| B.1.427 / B.1429 (Epsilon) | 2.1 |
| C.37 (Lambda) | 3.4 |
| B.1.621 (Mu) | 13.3 |
| B.1.617.1 | 5.35 |
| B.1.617.2+ (Delta + E484K) | 9.5 |

To obtain Omicron sequences for immune prediction, we download Spike sequences from a composite FASTA file of the whole genome nucleotide sequence of sequences available on GISAID and identified with a “B.1.1.529” Pangolin lineage, has been identified as the Omicron variant of concern by WHO. (By the time of submission of this paper, additional lineages have been differentiated but also identified as Omicron, and it is now possible to identify all Omicron sequences in the database.) The Spike gene nucleotide sequence from the annotated NCBI Reference Sequence (GenBank record NC-045512) is available for download from NCBI at https://www.ncbi.nlm.nih.gov/sars-cov-2/. [116] We perform pairwise alignments between each individual Omicron nucleotide sequence with the Spike nucleotide sequence from the reference genome [107] using the local pairwise Striped Smith-Waterman (SSW) method as described above (but with nucleotide rather than amino acid sequence). The region of best alignment sequence can then be translated. We exclude any alignment that produced degenerate sequences (i.e., which would include an “X” or other ambiguous amino acid character upon translation). Each resulting Omicron Spike protein sequence is then processed in the same manner as the “aligned” sequences described above, resulting in sequence in which all indels are positioned with respect to the reference protein. A Python (Google Colab) notebook including the preprocessing steps performed for Omicron variant sequence data is available for download at https://github.com/bahrad/Covid/blob/main/Covid_Predict_Omicron_Resistance.ipynb.

### Model Architecture

Fig. 1 shows an overview of our deep neural network model architecture. In this paper, the input is a protein sequence of length 1273 where we are analyzing exclusively SARS-CoV-2 sequences, when we analyze multiple coronavirus species we set the length at 1500 and pad or truncate input sequences. We tokenize each amino acid, and the deletion symbol “-”, to a distinct nonzero integer. We assign a to a nonzero integer as well. A position with the symbol “*”, which was padded as part of the alignment process described above, is assigned a zero token. Likewise, ambiguous amino acids with symbols X, B, J, or Z are assigned to zero. All positions with a token value of zero are masked. The token sequence is then transformed a matrix of embeddings of each token in the input sequence, in which the embedding includes both the token and its positional information. The embedding begins with a random initialization, and its parameters are then allowed to learn during the learning task.

We employ a transformer architecture for sequence encoding, which was first described in developing an encoder-decoder architecture for machine translation of text. [117] The transformer architecture is a modular multi-head structure, in which each head consists of an attention layer and feed forward neural network, wherein the head outputs are added and normalized to provide a sequence encoding. [117] The token and position embedding dimension (*N*_*E*_) that we use are large, because the sequence lengths are either 1273 or 1500 and the embedding for transformer-based architectures includes positional as well as token information. Consequently, we use a convolutional neural network (CNN) layer with a kernel width of 1 and *N*_*C*_ filters before the transformer as a way to reduce the required size of the transformer heads in order to allow us to load our model on a single processing unit for efficient computation.

We then use the encoded sequences to do classification or regression, depending on the learning task. We add two layers which assist in classification but also allow us to visualize, and potentially interpret the models that we train. The first is a self-attention layer following the structure inspired by [96] and which we utilized in [100] in conjunction with a Bidirectional Long Short Term Memory (Bi-LSTM) encoder. Here, we apply this structure to be able to readily access sequence-level classification, since the attention heads cannot individually, nor in sum, provide full attention visualization across the sequence (see, e.g., [103]). The second is an intermediate *N*_*H*_-dimension densely connected layer, which can project an *N*_*H*_-dimensional embedding of the sequence as a whole. In our studies of patient status metadata classification, we allow for the input of integer-valued demographic variables to the *N*_*H*_-dimensional embedding layer. As explained above, we use integers for age and sample collection date, and a 0/1 value for gender. These embeddings are then fed to an output layer. The output layer can be either of the following: (1) A single node with sigmoid activation for either binary patient clinical status classification or the regression used to predict the Omicron variant’s reduction in neutralizing antibody activity. (2) A clipped linear output restricted between 0 and 1 for regression to sample date. (3) A softmax layer with four nodes for the coronavirus genus-level taxonomic classifier.

For the studies shown herein, we sought to use the same hyperparameters across all models, as the input data was of the same form: approximately the same length (1273 or 1500) of tokenized amino acids. We employ 8 heads, a feedforward network (FFN) of 64 dimensions, and a dropout of 0.1 for the transformer layer. While differences were not substantial, this was the optimal combination of values we found for a cross-validation of the training data used for the patient metadata study. To confirm, we evaluated combinations of varying the number of heads between 4, 6, and 8; the number of nodes in the FFN were set at 32, 64, and 96 nodes; transformer dropout between 0.0, 0.1, and 0.2; N_C_ of the CNN at 300, 400, and 500; *N*_*H*_ at 64 or 128; and *N*_*E*_ of 1000, 1200, 1500, and 1800. We also found that varying parameters did not result in a substantial difference in model predictions in the other classification and regression tasks shown in this paper. Model training was done using the standard Adam gradient descent fitting algorithm in Tensorflow 2, with binary cross-entropy and mean squared error as the loss functions for classification and regression tasks respectively. The learning rate parameter set at 1× 10^−4^, after evaluating rates of 5 ×10^−6^, 1 × 10^−5^, 1 × 10^−3^, 5 × 10^−3^, 1 × 10^−2^, and 1 × 10^−1^.

We select the number of epochs for training based on our observation that, in general, the number of epochs required for classification tasks is much lower than that for regression tasks. In the case of binary classification for patient metadata, we train for 75 epochs, after evaluating epoch ranges from 50 to 200. We do not use early stopping with a validation set, as found that reduces the training data and causes underfitting, and instead early stopping is set at 20 epochs with no net change in the binary cross-entropy loss. For the regression to sample date task, we set the limit at 600 epochs, with an early stopping time set at 200 epochs. For the taxonomic classification task, we train for 25 epochs, with an early stopping condition set based on no change in the loss of a validation set, randomly selected from 20% of the samples in the training data, for at least 5 epochs. Finally, for the Omicron immune evasion prediction task, we set the maximum number of epochs at 200, employing early stopping after 10 epochs in which the mean squared error loss of a random 10% of training samples no longer declines. For all of the neural network training tasks, we used multiple runs to evaluate run-to-run consistency given the randomization of initial weights and other inherent randomization of training.

As further described in the Results & Discussion, we also benchmarked the transformer-based model against eXtreme Gradient Boosting (XGBoost), a decision tree-based ensemble learning method [118]. For XGBoost, after hypertuning, the following parameter values were used for XGBoost: subsample rate of 0.8, lambda regularization of 1.0, maximum depth of 10, learning rate of 0.001, gamma of 0.0, column sample by tree of 0.8, and the GPU-optimized predictor. XGBoost is implemented using the Python package in xgboost 1.5.1 in the Google Colab GPU runtime environment. Because XGBoost results may also change due to randomization, we evaluate the run-to-run consistency of XBoost under different random number generator seeds. We generally found that the results shown in this paper were consistent with negligible standard deviation between runs on the same data and with the same parameters.

For the purposes of model analysis and interpretation, we look at two scales: First, *attention*, which shows where the model attends along the sequence (i.e., amino acid position in a protein sequence) and can identify patterns at the sequence level. [100], [101] Second, *embeddings*, in which the *N*_*H*_-dimensional encoding of a sequence at the layer shown in Fig. 1 (see above) is obtained for a trained model, and then the embeddings of a group of sequences post-training are visualized or clustered to find patterns at the group level. [119] We have previously shown that, for example, we can reveal underlying taxonomic categorization when we visualize the embeddings of 16s rRNA sequences from a model that has been trained to identify disease status. [100]

### Implementation

Model training was done using the standard model fitting algorithm provided by the Keras API on top of Tensorflow 2, with. The hardware used for neural network model training and evaluation, as well as the data pre-processing described above, consists of Nvidia Tesla P80 GPUs (primarily) and Google Cloud Tensor Processor Units (TPUs) the Google’s Colab environment, running Tensorflow 2.70 and Python 3.7.12, and Nvidia Tesla V100-SXM2 GPUs on the Drexel University Research Computing Facility (URCF), running Tensorflow 2.4 and Python 3.8. Because model hyperparameters were not varied between tasks, training times per epoch are a function of the training data set size across all tasks and whether we used a TPU or GPU for training. For example, patient metadata classification trains on 44,003 samples, which requires 51 sec/epoch in the Google TPU environment had 51 sec/epoch, while on a GPU unit in the URCF, the time per epoch was 480 seconds, representing a 9.2-fold TPU-speedup. By contrast, the model to predict Omicron neutralizing antibody activity classification trains on 2,245 samples, and it requires 4 sec/epoch on the Google TPU.

## RESULTS & DISCUSSION

### Regression Based on Sample Collection Date

To validate the interpretability of our deep sequence modeling approach, we demonstrate that embeddings and attention can be used to visualize the results of our model’s training in an exemplary machine learning task. Here, we show the results of regression to the sample collection date of sequences in a database of raw, unprocessed spike protein sequences available on GISAID as of October 1, 2021. For this task, we analyze the earliest sample collection date included in the GISAID database for each distinct sequence in the data set. The training data set consists of sequences that are found more than four times in the database. The validation set consists of sequences found only two or three times in the database. Notably, some sequences are found very frequently, such as certain sequences in the B.1.1.7 (Alpha) and B.1.617.2 (Delta) lineages. For the purpose of this task, these sequences are included once and weighted equally to other sequences. Trivially, the regression should “predict” that a sequence will have appeared at the same time as others within the same lineages, as the lineages are defined based on sequence similarities at particular sites with the SARS-CoV-2 genome. In particular, the lineages, following the Pango scheme [59] are based in large part on changes in the spike protein sequence, which should be reflected in our model, which is trained on spike protein sequence data. Accordingly, trends we see in the embedding and attention should reflect the classification.

As an initial matter, we confirmed that the trained model’s error in the prediction data set were within a reasonable error margin. To do so, we normalized the square root of the mean squared prediction error (RMSE) on validation set data by the RMSE of simply predicting each label with the overall mean value of all labels. We found that the RMSE of the trained model was 0.0962 and normalized RMSE was 0.534. By comparison, the normalized RMSE on the training data set was found to be 0.37. Therefore, while the RMSE increased when generalizing the model to the validation set, it remained significantly below 1.0, indicating that the model was predicting significantly better than chance.

Fig. 2 further demonstrates the model’s performance by showing that a t-SNE (t-distributed stochastic neighbor embedding) plot [120] of the *N*_*H*_-dimensional (*N*_*H*_ = 64) embeddings of sequence samples in the validation data set. Fig. 2 is consistent with the intuition described above: A lineage will generally consist of sequences that emerge at a common point in time. As such, modeling sequences by the date of their first emergence groups the sequence embeddings from the same lineages together and separates them from embeddings of other lineages. Notably, these samples are from a validation data set which includes raw sequence data, i.e., they include ambiguous residue calls and are of all different sizes. Assuming that all sequences between 1270 and 1273 amino acids in length are “normal” (i.e., are the original length of 1273 amino acids or contain known common deletions), then 2,619 sequences of the 58,747 sequences are of nonstandard lengths (634 are fragments with a length less than 500 amino acids). Moreover, a majority of sequence – 39,008 – include one or more ambiguous amino acid calls. These issues may account for instances where the sequence embeddings do not group by lineage as expected. The overlap between B.1.617.2 (Delta) and AY.4 is also expected, as AY.4 lineages (e.g., AY.4.2) are recognized as Delta sublineages which have emerged during the Delta outbreak. [121] In sum, Fig. 2 shows that, without doing 1) further alignment, 2) phylogenetic tree-building, or 3) classification based on sequence polymorphisms, we can replicate SARS-CoV-2 lineages and predict the date of emergence of a sequence.

**Figure 2.**
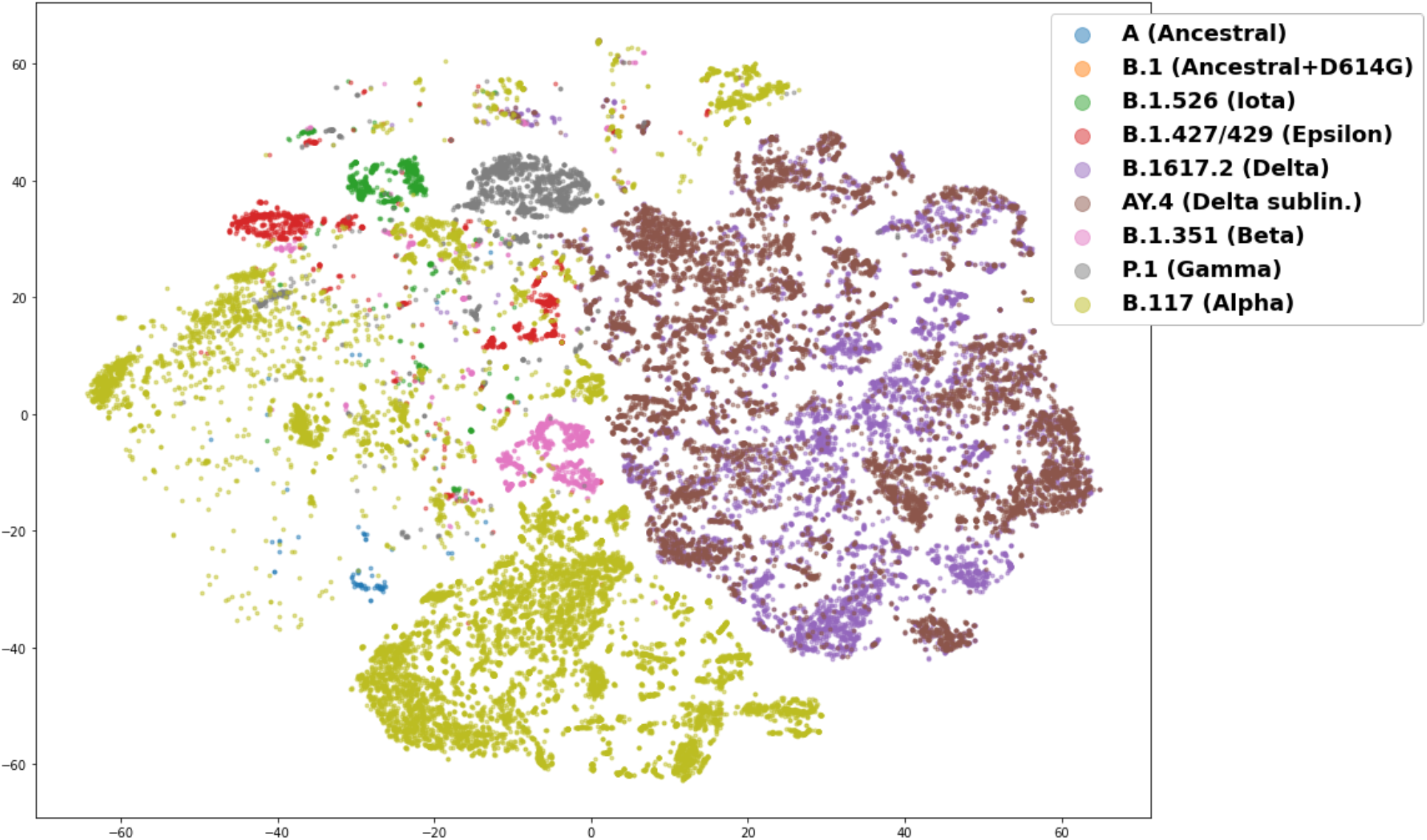
Plot of a two-component t-SNE analysis (with perplexity of 50) for the 64-dimensional embeddings of raw spike protein sequence data following a prediction task for the date of first sample collection (in days since December 1, 2019) of a particular sequence. The training data consisted of sequences found 4 or more times in the global data set (all spike protein sequences available on GISAID as of October 1, 2021), and the validation data shown here consists only of raw, unprocessed spike sequences found 2 or 3 times in the global data set. The lineages shown in this plot are provided in the legend and consist of the ancestral lineage (A), as well as as variants of concern and interest including Alpha (B.1.1.7), Beta (B.1.35.1), Iota (B.1.526), Gamma (P.1), Epsilon (B.1.427/429) and Delta (B.1.617.2), and AY.4, a Delta sublineage. Many lineages can be clustered with the embeddings; Delta and the AY.4 Delta sublineages cluster into a large but well separated group, while Alpha is spread out.

Visualizing attention further supports our model. To analyze attention, we show the results of modeling the aligned data set, in which sequences were aligned to the reference sequence and padded with mask characters for an equal length of 1273 amino acids. This was done to better show consistent attention values across sequences in the validation data set which are of different lengths. We did not correct for mask or noise symbols in the spike sequences. The prediction errors were in line with the raw sequence data set, with a RMSE of 0.0970 and normalized RMSE of 0.53. Fig. 3 shows the mean attention for each sequence in the validation data set (sequences occurring more than 2 or 3 times in the global October 1, 2021 data set) binned by date of first emergence. We can see in Fig. 3 that attention values change as the pandemic progresses, which corresponds with the introduction of new variations in the spike protein sequence over time. Fig. 4A demonstrates how major lineages have emerged and retreated during the time periods corresponding to the binned dates in Fig. 3.

**Figure 3.**
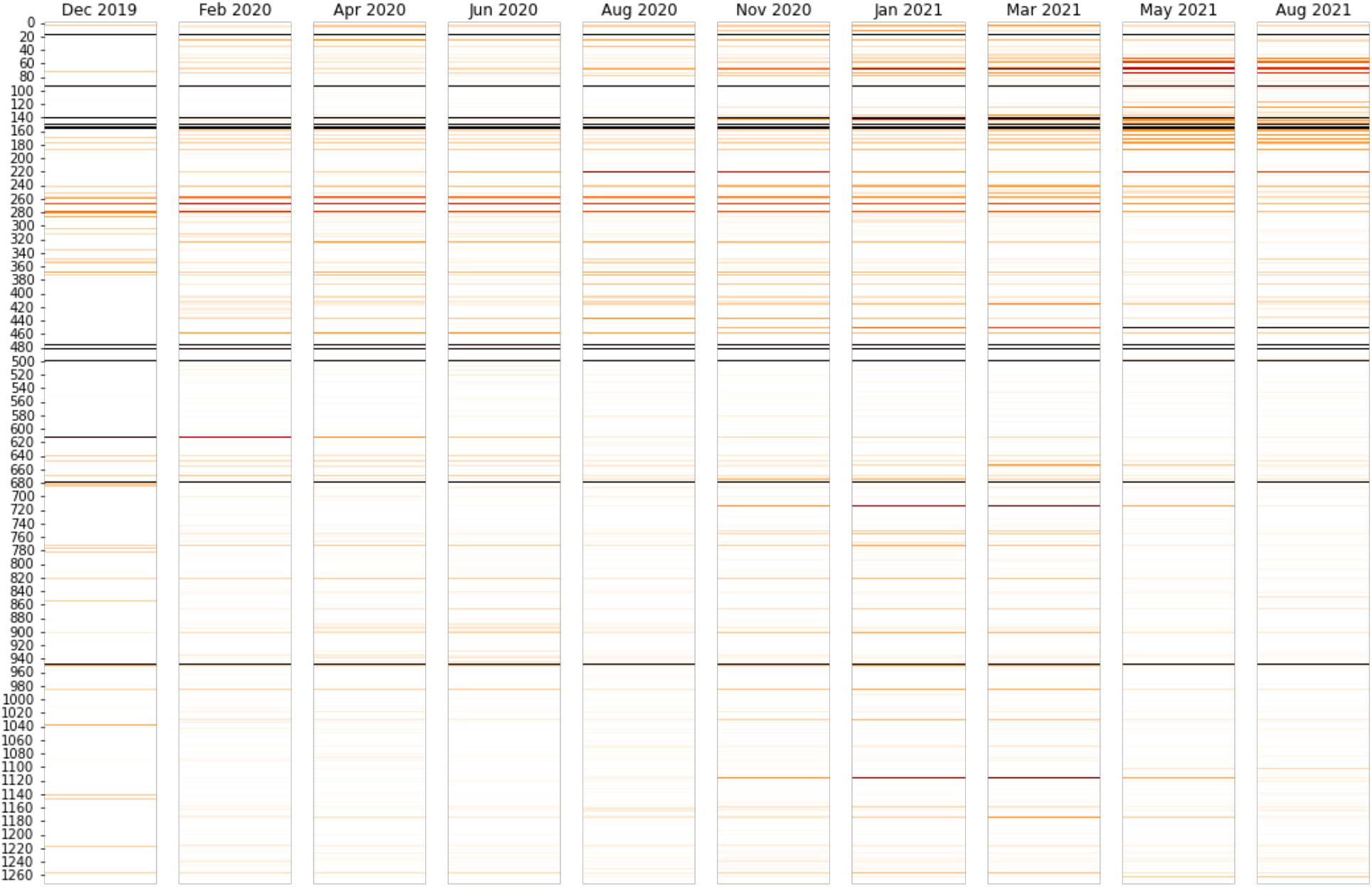
Mean attention at each amino acid position for sequences in the validation data set (aligned sequences occurring 2 or 3 times in the October 1, 2021 global data set) categorized into bins based on the date of first emergence. The intensity of the color attention is log-normalized by the maximum attention for all sequence positions, so that the peak values are black and intermediate attention levels are various shades of red. The mean attention values vary over time as waves of new viruses emerge. For example, sites between 600 and 1200 become important for the emergence of Alpha in the winter of 2020-21. Some specific positions, e.g., 478 and 681, have high attention throughout time because of their changes from early in evolution, Alpha, to later, Deltam while other sites, such as 452, which is characteristic of Delta, become important as that variant emerges.

**Figure 4.**
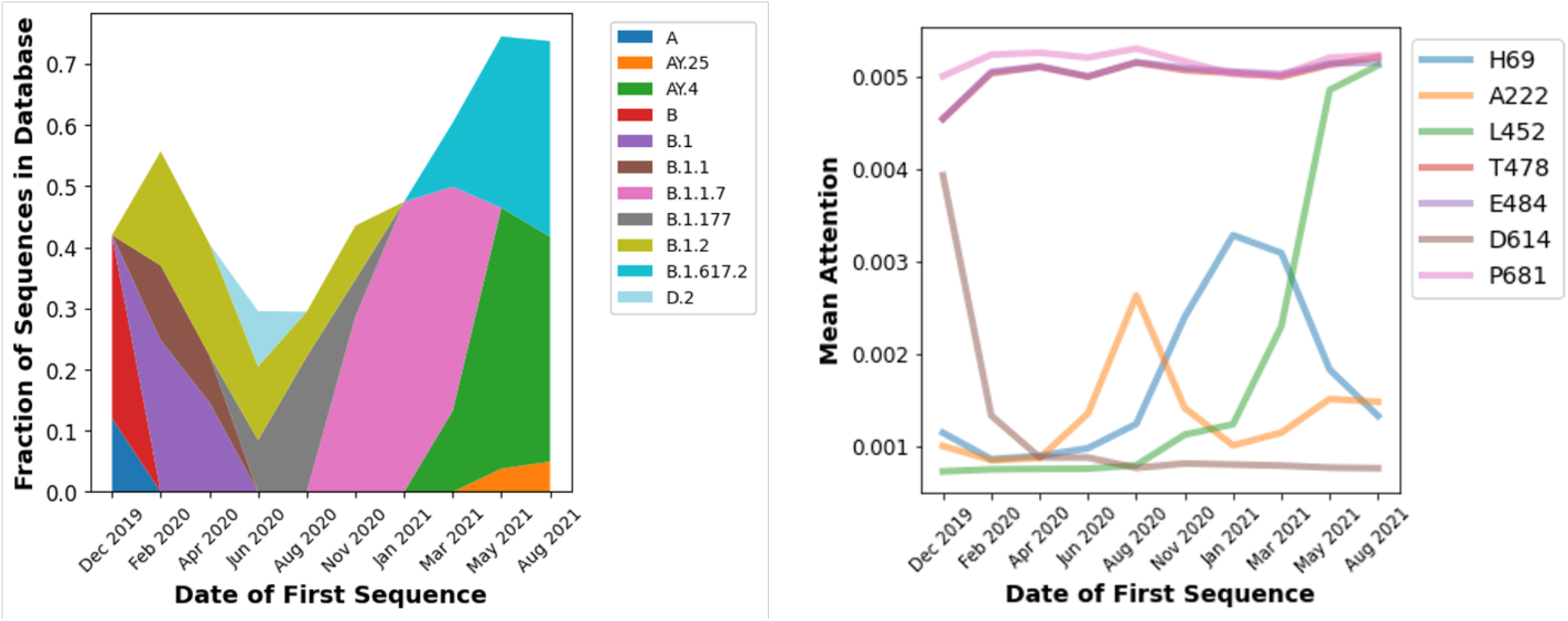
**(A – Left)** Relative prevalence of the three most frequently observed lineages (by sequence) in the October 1, 2021 data set in each time period identified in Figure 3. Because less frequently observed lineages are excluded for clarity, the values sum to less than 100%. **(B – Right)** Mean attention levels for exemplary protein sequence positions with date of first emergence in time periods corresponding to Figure 3. The legend shows the amino acid in the reference (wild type) sequence for each position.

Comparing Figs. 3 and 4A, we can see how attention levels in the Winter of 2020 and Spring of 2021 at various positions increase relative to earlier time points, and in some cases decrease later in 2021. Broadly, the attention patterns correspond to the increase and decrease in prevalence of lineages in Fig. 4A, in particular the B.1.1.7 (Alpha) wave, which succeeded earlier lineages that originated in China and then spread to Europe and North America, and which was followed by a wave of Delta (B.1.617.2) and sublineages. Fig. 4B shows the patterns in greater detail for exemplary sequence positions. For example, a characteristic Delta mutation, L452R, increases as 2021 continues and Delta emerged. [122] By contrast, the attention at site 614 is highest earlier in the pandemic. This corresponds to D614G being the first major mutation to emerge after the pandemic began, and then subsequently becoming a ubiquitous aspect of all subsequent lineages. [123] Consistent with the rise and fall of the prevalence of B.1.1.7 shown in Fig. 4A, Fig. 4B shows that the rise and fall of the attention level at position 69. Positions 69/70 are where deletions occur in the B.1.1.7 (Alpha) lineage, which have not been found in Delta and Delta sublineages which began to dominate later in 2021. We see a more complicated picture for position 222, as the A222V mutation emerged late in 2021 in the Delta AY.4.2 sublineage. [121] The A222V mutation, however, had also been found in mid-2020, which corresponds to the earlier peak in Fig. 4B. [124] The reason why some attention values at certain sites remain consistently high over time could be because there consistently has been some substantial level mutations at those sites throughout the pandemic. For example, the attention at positions 478 and 484 track each other at high levels and coincide on the same track on the plot. This may be explicable because position 478 is in the receptor binding domain (RBD) of the spike protein, and it had a high level of mutations in summer of 2020 [125] continuing through later in 2021 when the T478K mutation emerged as a characteristic mutation of the Delta lineage. [122] Similarly E484Q has been a characteristic mutation in the Delta lineage, but E484K has been an aspect of other pre-Delta, such as Beta and some Alpha sublineages. [126]–[128] Similarly, P681H was found in the highly prevalent Alpha lineage [129], and P681R was found in the Kappa and then later Delta lineage. [130] Accordingly, it may be the case that a consistently high attention suggests that the model pays attention to the site because it was not a point of variation at early time points. The attention does vary though in subsequent time periods where it is found in some important lineages (such as Kappa and then Delta) but no other highly prevalent lineages (e.g., Alpha). The model thus needs to use that site to predict the emergence date of sequences across all time periods.

### Predicting the Severity of Patient Outcomes

As detailed in the Methods section, we utilize GISAID samples with patient metadata to develop a model for the relationship between severity of COVID-19 disease and the spike protein sequence. Approximately 147,000 of the 3.9 million samples available on the October 2021 cutoff date for inclusion in our study include any patient metadata as well as readable spike protein sequence information. Many of the metadata entries are simply “unknown” or not applicable to patient status, e.g., include information about sample collection. Additionally, as described in the Methods and shown in Supplemental Table 1, many descriptions cannot be used together in a coherent classification task. For example, we could compare patients who are alive and dead, but the patients who are not listed as being dead would have a wide range of outcomes from asymptomatic to ICU admission. Accordingly, we focused on metadata that could be assigned to two broad categories, “Mild” (0) or “Severe” (1) following the scheme in Supplemental Table 1, based on the NIH Clinical Guidelines for COVID-19. [108] After eliminating samples which cannot be classified as Mild or Severe, 54,081 samples remain.

#### Predicting patient outcomes using machine learning models based on both sequence and demographic information

When developing a model to link sequence to disease outcomes, we need to also consider demographic variables. Throughout the pandemic, there has been consistent epidemiological and clinical evidence that age and sex/gender (male) are major risk factors for more severe COVID-19 symptoms. [131]–[133] Earlier studies have shown that GISAID data also supported a correlation between the demographic metadata which are available—age and gender—to clinical severity. [76], [77] Figs. 5A and 5B show the relationship between these variables and clinical severity, which show, as expected, a correlation with age (increasing age results in increasing mean severity) and sex/gender (male patients have a greater prevalence of severe outcomes). Notably, the age data do not show a consistent trend at the extremes of young and old. Extreme age values, however, are represented by far fewer samples in the data set, and thus may be susceptible to study bias of which samples were sequenced. For example, if more infants (with sequenced samples in the GISAID database) were hospitalized, even for incidental reasons, that would be reflected as a more severe case. Or, if very old patients were sequenced as part of a study of elderly patients who had survived or had a milder course of disease of disease than expected for their age, that would skew a small number of samples milder.

**Figure 5.**
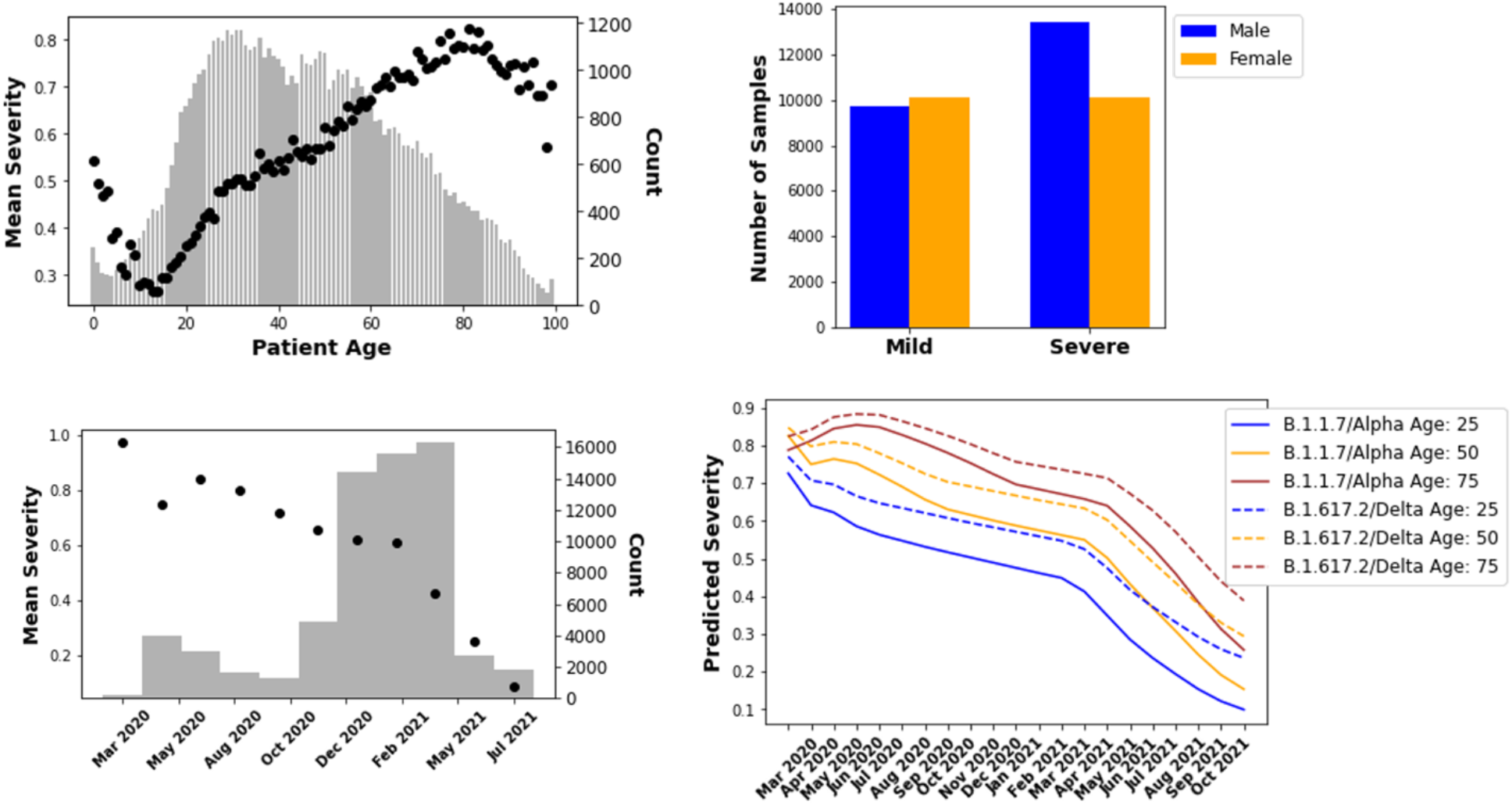
(**A–Upper Left**) Mean of clinical severity, where 0 is Mild and 1 is Severe (i.e., probability of a severe case) by patient age in the GISAID database, where age metadata are available. The bars show the count of samples for each age. (**B–Upper Right**) Number of mild and severe cases for each sex/gender as identified in the “gender” patient metadata field for GISAID samples. (**C–Lower Left**) Mean clinical severity (probability of severe case) by sample collection date recorded in the GISAID data. For clarity, data have been binned over time periods; the number of samples in each bin is shown by the bars. (**D–Lower Right**) Model predictions, including sequence, age, date, and gender information, of mean severity over time of patient samples with sequences from Alpha (B.1.1.7) and Delta (B.1.617.2) lineages. Three different ages are run, as indicated in the legend, and the gender variable is set to male. The predictions shown here are the averages for the 30 most prevalent sequences (frequency found in database) for each lineage. We can obtain a prediction over time of variant severity, accounting for time in pandemic and age of patient.

Besides patient demographics, we find a substantial trend in the frequency of severe outcomes based on sample collection date. Fig. 5C shows how the number of samples with severe disease in the GISAID database drops as a function of time through the pandemic. There has been a particularly sharp decrease since approximately February 2021. This trend has been consistent even as the prevalent genetic compositions of the virus have changed in many different ways over time. We can see this in Table 2, which shows the percentage of severe cases, mean age, and percentage of male patients for the patient metadata validation set obtained as detailed in the Methods section. For example, the B.1 lineage (D614G) was first detected January 1, 2020 (https://cov-lineages.org/lineage_list.html), and, as Fig. 4A shows, was no longer highly prevalent by the mid-2020. In the validation data set, 73% of samples indicated a severe outcome for the B.1 variant, the ancestral genome with a D614G mutation. By contrast, only 60% of patient outcomes are severe for Alpha (B.1.1.7) sequences, and that drops to 42% for Delta (B.1.617.2) and further to 26% for AY.4 Delta sublineages. Although the average age is lower for Delta patients, that is insufficient to explain this drop, which is seen across sequences with higher ages as well. Overall, the results shown in Table 2 contradict repeated studies showing that Alpha resulted in elevated hospitalizations, ICU admissions, and other markers of severe outcomes as compared to ancestral lineages, that Delta was yet more severe, and that overall other variants of concern emerging later in the pandemic lead to more severe outcomes overall. [134]–[140]

**Table 2.** Mean age, percentage of samples which are from patients with severe COVID-19 disease, percentage of samples from male populations, and the mean sample collection date of samples in the GISAID patient metadata validation data set from the lineages that appeared at least 400 times (i.e., frequency > 400). Mean sample collection date is defined in terms of days since December 1, 2019. The most recent lineages (highest sample date) trend to lower severity.

| Lineage | Age | % Severe | % Male | Sample Date | Frequency |
| --- | --- | --- | --- | --- | --- |
| AY.4 | 39.4 | 26% | 53% | 631 | 817 |
| B.1 | 49.8 | 73% | 42% | 314 | 994 |
| B.1.1 | 50.3 | 70% | 49% | 314 | 696 |
| B.1.1.519 | 49.9 | 68% | 47% | 485 | 1583 |
| B.1.1.7 | 48.3 | 60% | 50% | 509 | 3298 |
| B.1.351 | 45.7 | 52% | 50% | 481 | 849 |
| B.1.617.2 | 40.0 | 42% | 48% | 604 | 3901 |
| P.1 | 48.3 | 64% | 48% | 548 | 877 |

Our model, however, cannot account for changes in how COVID-19 is treated and can be prevented. In particular, we are unable to control GISAID data for vaccination, as there is still very limited information about vaccination for samples in GISAID metadata (at least as of November 2021). The early decline in 2020 in average case severity is consistent from a Canadian study showing that the case fatality rate (CFR) decreased between the first and second waves in Ontario prior to any vaccination, even when controlling for age and increased testing. [141] The latter decline may be accounted for by better understanding of how to treat COVID-19, and, later in 2020, the emergence of monoclonal antibody therapies. The accelerated decline in case severity starting in early 2021 is likely attributable to vaccination, particularly of older populations, which would also be consistent with the lower mean ages of Delta and its sublineages as indicated in Table 2. Countries in which vaccines were widely available among the elderly in early 2021 and general population through 2021 are highly overrepresented in GISAID sequence data: of all sequences in GISAID through our October 2021 cutoff, 55% were in Europe and 34% were from North America. The overrepresentation is less acute in the subset with patient metadata, but still 69% of the GISAID sequences with metadata for patient status were from Europe and North America combined.

In addition to treatment and vaccination effects, the sequence data are also impacted by artifacts arising from the context of where sequences are obtained. Early in the pandemic, we observe that every sample is from a hospitalized patient, which is consistent with limited testing and a bias towards hospitalizing COVID-19 patients to isolate and treat them. Later sequence samples are likelier to come from population screening studies, where subjects could be asymptomatic or only mildly symptomatic. It may even be that the “real” average disease severity has decreased even faster than our analysis reflects. We categorize hospitalizations as “severe” cases, acknowledging that some fraction of those cases will not be in fact severe in those cases where GISAID metadata cannot distinguish whether the patients was admitted for other reasons but are not severely ill from COVID-19 itself. There is evidence that the fraction of incidentally hospitalized cases may be increasing over time due to vaccination in the United States and other countries. [142]–[145] The reason proposed for why vaccination would increase incidental hospitalization of non-severe cases is that more patients being admitted for other reasons will test positive as they have asymptomatic or barely symptomatic (paucisymptomatic) cases. Moreover, the foregoing biases could explain why we observe an overall high proportion of severe cases in our analysis.

Accordingly, we include sample collection date as a demographic variable in training the neural network, along with age and gender. As shown in Fig. 1, these variables are fed into the neural network model at the *N*_*H*_-dimensional densely connected layer. A sample’s demographic variables are thus embedded along with the spike protein sequence in the *N*_*H*_-dimensional embedding vectors obtained from that layer. Table 3 shows the classification metrics of the Transformer-based neural network model and XGBoost on the validation data set. The training and validation sets are constructed to avoid sequence overlaps, as detailed in the Methods section. We compare the results of our neural network model to XGBoost, as it is a decision tree-based ensemble learning approach that has proven to be useful in biological sequence machine learning applications as it is 1) fast, 2) highly effective at classification, 3) robust to missing values (which frequently arise in GISAID data due to missing or ambiguous amino acid codes), and 4) can provide a ranking of feature importance, which we use to validate the interpretability of our neural network model below. [146], [147]

**Table 3.**
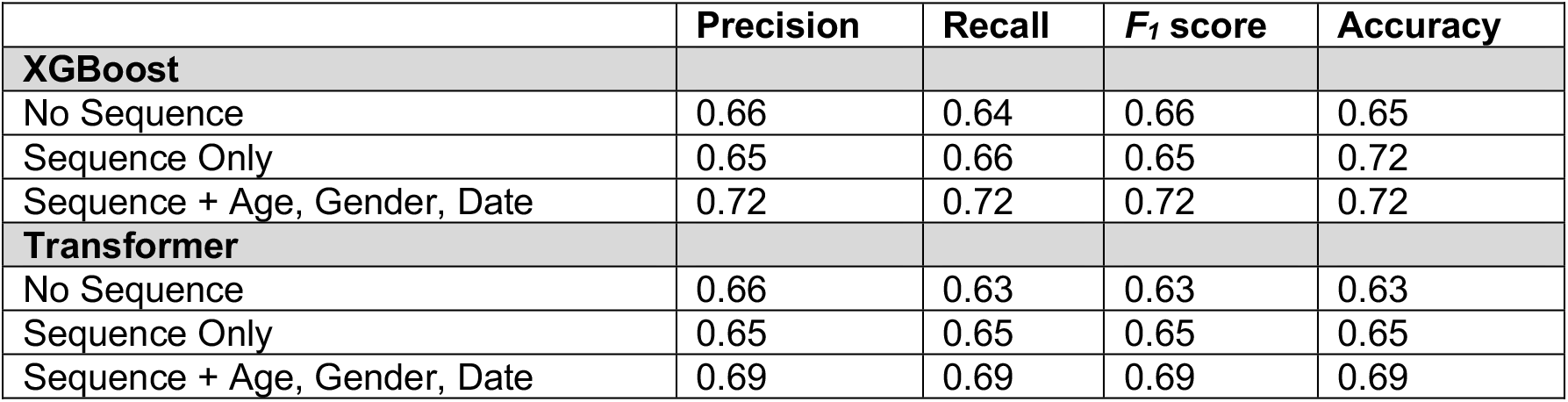
Classification metrics for the patient metadata validation set for the neural network model of Figure 1 and XGBoost model trained on individual samples (rather than sequences). Prediction results shown are based on training on age, gender, and date as the only independent variables, sequence alone, and the sequence combined with other variables. There 8,948 and 12,965 samples of Mild (Class 0) and Severe (Class 0) patient outcomes; shown here are average metrics weighted to account for the class imbalance. Classification accuracy of the Transformer-based neural network model approaches 70% and is only somewhat lower than XGBoost.

| | Precision | Recall | $F_1$ score | Accuracy |
| --- | --- | --- | --- | --- |
| **XGBoost** |  |  |  |  |
| No Sequence | 0.66 | 0.64 | 0.66 | 0.65 |
| Sequence Only | 0.65 | 0.66 | 0.65 | 0.72 |
| Sequence + Age, Gender, Date | 0.72 | 0.72 | 0.72 | 0.72 |
| **Transformer** |  |  |  |  |
| No Sequence | 0.66 | 0.63 | 0.63 | 0.63 |
| Sequence Only | 0.65 | 0.65 | 0.65 | 0.65 |
| Sequence + Age, Gender, Date | 0.69 | 0.69 | 0.69 | 0.69 |

Overall, as Table 3 shows, both our neural network and XGBoost are able to correctly predict a substantial majority of both the mild and severe cases. We also found that the XGBoost does outperform the neural network model. This was as expected, since most of the differences between spike protein sequences, particularly at early time points in the pandemic are at specific sequence positions where XGBoost should perform best. The neural network model was found to be able to consistently make nearly as accurate predictions. Moreover, for both methods, including sequence as well as the demographic variables led to a superior performance than considering each alone.

The combined sequence and demographic model can also be qualitatively validated. Fig. 5D shows predictions made by training the Transformer-based neural network model that using sequence as well as age, gender, and sample collection date. We simulated the trained model for dates starting in March 2020, well before either Alpha (early 2021) and then later Delta (mid-2021) were prevalent and their sequences had emerged (September 2020 and December 2020 respectively). [148], [149] As expected, the model predicts that Alpha and Delta cases would have been more severe had they occurred earlier in the pandemic. Moreover, the model predicts that, overall, Delta cases would be more severe than Alpha, in agreement with the above-cited clinical and epidemiological literature. In sum, once we incorporate sample date in our model, we are able to produce predictions that are validated both quantitatively and qualitatively.

We note that we are limited in the demographic information that we can include given that the sequence data on GISAID does not have additional information. As a result, our model cannot account in other important factors that increase the risk of severe COVID-19, including comorbidities and racial disparities. [150] Also, patient metadata may be snapshots in time. For example, a mild case may evolve into a severe case without updating its annotation. We also do not consider the geographic origin of cases. The severity of cases may be related to different countries and regions’ enforcement of non-pharmaceutical interventions (NPI), the differences within a country in who has a large number of contacts, which may itself change over time, level of circulating virus, and differences in hospital capacity and standards of care. [151]–[153]. However, there is a strong confounding effect between sequences and countries. [65] As a result, the relationship between sequence and severity will in fact reflect regional-specific factors to some extent, particularly for sequences that do not become widespread in multiple regions. However, given the small fraction of sequences for which patient metadata are available, there are too few sequenced samples in individual countries or regions to enable machine learning based on geographic variables.

#### Interpreting the disease severity model by visualizing embeddings and attention

To visualize sequence embeddings, we select a random equal sampling of the 45 most prevalent lineages. Sampling allows the visualization to be more tractable while retaining the structure of the data, i.e., to avoid having lineages that have many more sequences in the database, particularly those from Alpha and Delta lineages, swamp the signal from other sequences. In Fig. 6, we show how the different input variables, sequence, age, gender, and sample collection date, influence the embedded structure of the validation data set. To visualize the effect of sequence on a sample’s embedding, we quantify the distance between the sample’s spike sequence and the spike reference sequence, which is the consensus of ancestral genomes. We use the mismatch frequency divided by length of the spike reference sequence as a per cent mismatch to measure sequence distance.

**Figure 6.**
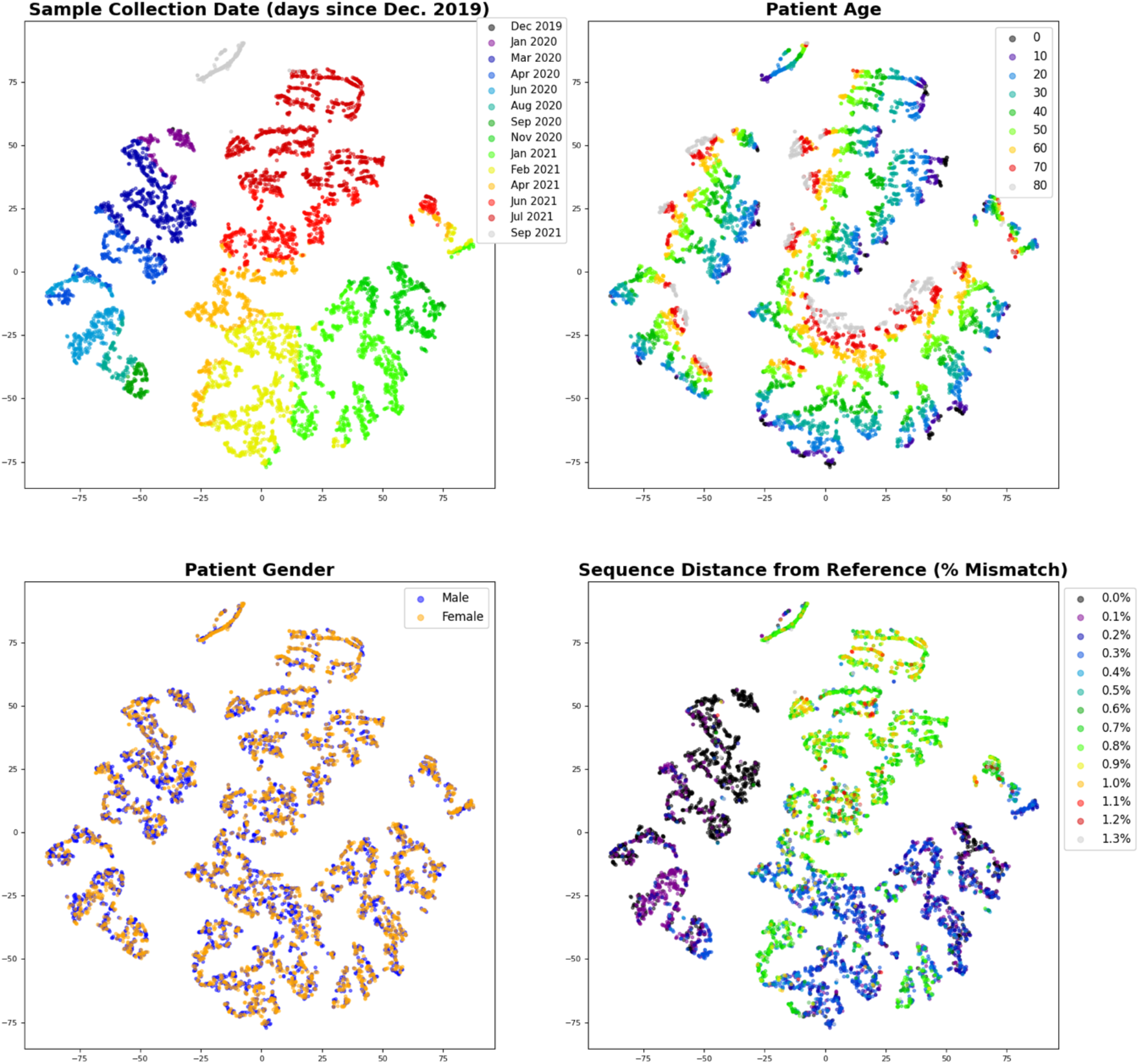
Sequence embeddings obtained from the trained neural network model and visualized based on a 2-component t-SNE (perplexity of 30). The plots all show the same t-SNE visualization of a random sampling of 150 samples each of the 45 most prevalent lineages in order to achieve sequence and sample date diversity. Clockwise starting with the upper left, the graphs show the t-SNE distribution labeled by sample collection date, patient age, sequence distance, and gender respectively. Variables are binned as indicated in the legend. Sequence distance is the number of mismatches between the sequence of the sample to the reference protein sequence divided by the reference protein sequence length, disregarding any padding or ambiguous amino acids in the sample sequence. The embeddings are heavily influenced by variant emergence and thus time, in addition to age. Gender has little effect.

We expect that the t-SNE plots of the embeddings in Fig. 6 should reflect how the model performs its ultimate regression. The *N*_*H*_-dimensional embedding layer is the layer immediately before the final sigmoid-activated node that outputs the class probability (i.e., probability of that a sample is from a severe case) with is rounded to the mild/severe (0/1) prediction. Therefore, at the *N*_*H*_-dimensional layer, embedded samples will be separated in the embedding space in such a manner that they can be most accurately classified based on the trained model parameters. The t-SNE plots should thus accentuate similarities and distances, the visualization should reflect the separation and clustering of the embeddings according to how they will be classified.

Fig. 6 shows, as expected, consistent patterns within the relative positions of sample embeddings according to age, sample date, and sequence distance. The exception is gender, which fails to show separation between male and female samples. The lack of a pattern for gender in Fig. 6 is consistent with Fig. 5B, which shows that the effect of gender on disease severity is not as significant as that for age or sample date. Severe cases account for 58% of samples from male patients, as opposed to 53% from female patients. (As noted above, the overall likelihood of severe cases in our GISAID-sourced data set is much higher than would be expected in the general population.) By contrast, age and sample date do play an important role. As Fig. 6 shows, the patterns of age and sample date are orthogonal to each other. The relationship between age and sample date patterns is consistent with Figs. 5A and 5C, which show that as age increases, severity increases, but as sample date increases, severity decreases. Fig. 6 also shows that sequence is playing a role in the model prediction. We represent the effect of sequence by categorizing samples according to their degree of difference (distance) from the reference sequence that was used for alignment, as described in the Methods. The sequence distance is measured by mismatch frequency normalized by the length of the reference sequence (1,273 amino acids). Fig. 6 shows a pattern of separation and clustering among samples by sequence distance that is clockwise similar to sample date as the two will be correlated as discussed above. In sum, we can visualize how the neural network model utilizes the input variables to train the model.

We can also gain insight into the model through attention. To demonstrate that the attention of a sequence position in our model can correspond to the importance of that position to classification, we compare the attention values that we found at different positions in the spike protein sequence to XGBoost feature importance scores. A trained XGBoost model can be analyzed to identify feature importance through various scores that are based on characteristics of boosted tree models, such as the number of times a feature is used to split trees, or the gain in score towards the objective function obtained by splitting trees based on a feature. [154], [155]

Table 4 shows a comparison of the XGBoost feature score (gain) and attention score for the highest attention sequence positions. The ranking of the XGBoost feature score is related to the rank of the highest attention sequence features. The highest-scoring features in the XGBoost model, however, were age, sample date, and gender. This is likely because XGBoost uses a greedy approach to branch decision trees in variables that will most rapidly lead to the optimal classification. [118] Therefore, we expect that XGBoost will consider gender as an important variable as it is varies consistently in the training samples, while samples may have very similar or even identical sequences. By contrast, as Fig. 6 shows, it is less important to our Transformer-based neural network model. The XGBoost feature scores in Table 4 are likewise much more skewed towards a smaller number of variants than the attention scores from the neural network model. Despite these differences, as Table 4 indicates, the 10 most important sequence features per XGBoost were found among the 22 highest mean attention positions, showing a concordance between the attention and XGBoost scores.

**Table 4.** Amino acid positions with the highest mean attention score, comparing the XGBoost feature importance ranking and score (obtained from the gain measure of feature importance). This table includes the 10 highest-scoring features identified by XGBoost, which appear within the 22 highest attention values.

| Attention Rank | Amino Acid Position | Mean Attention Score | XGBoost Rank | XGBoost Score |
| --- | --- | --- | --- | --- |
| 1 | 1258 | 5.42 | 73 | 163 |
| 2 | 732 | 5.1 | 10 | 2090 |
| 3 | 142 | 4.96 | 1 | 6421 |
| 4 | 501 | 3.79 | 8 | 2272 |
| 5 | 69 | 2.14 | 5 | 2813 |
| 6 | 70 | 2.13 | 31 | 573 |
| 7 | 716 | 2.08 | 25 | 756 |
| 8 | 570 | 2.06 | 27 | 709 |
| 9 | 19 | 1.99 | 26 | 720 |
| 10 | 950 | 1.92 | 17 | 1352 |
| 11 | 144 | 1.34 | 6 | 2306 |
| 12 | 95 | 1.25 | 2 | 4698 |
| 13 | 222 | 1.07 | 4 | 3474 |
| 14 | 417 | 1.06 | 24 | 757 |
| 15 | 655 | 1.01 | 62 | 225 |
| 16 | 614 | 0.94 | 7 | 2288 |
| 17 | 1104 | 0.91 | 9 | 2240 |
| 18 | 241 | 0.87 | 134 | 78 |
| 19 | 243 | 0.86 | 335 | 7 |
| 20 | 242 | 0.86 | 250 | 23 |
| 21 | 477 | 0.86 | 14 | 1488 |
| 22 | 681 | 0.84 | 3 | 3559 |
| 23 | 1074 | 0.83 | 22 | 962 |
| 24 | 97 | 0.81 | 18 | 1168 |

Many of the high attention positions coincide with sites that have been identified as having a potential significance for virulence, infectivity, and/or immune evasion. For example, at the third-highest position, protein structure analysis suggests that G142D, which is present in some Delta and Delta sub-lineage sequences, may be linked to higher infectivity and immune evasion. [156] In the GISAID data, sequences containing G142D are in fact generally associated with lower severity, although that is partly a function of its more recent emergence which would result in a lower apparent severity for the reasons discussed above. The highly ranked sites also include locations known to be significant for differentiating SARS-CoV-2 variants, such as positions 69 (corresponding to the 68-69 deletion), 501 (where the N501Y mutation occurs, which is characteristic of B.1.1.7/Alpha) and 681 (P681R and P681H) as shown in Fig. 4B and discussed in the accompanying text.

The site with the highest mean attention, position 1258, however, has not been as well characterized. The most common mutation found in our training and validation data at that site was E1258D, although we also found E1258H. E1258Q has been observed before as well, and the site has been linked to coincident mutations that may result in a spike protein missing its terminal region. [157] It appears that these spike mutants would cause the spike protein to accumulate in the plasma membrane, which would result in syncytia, large multinuclear cellular masses, which may result in heightened virulence. [157] We did not observe such truncated spike proteins in our patient data set. However, we do see that E1258D is associated with much higher disease severity. Among the 2.6% of samples with an E1258D mutation, 99% were categorized as severe cases. (Our model correctly predicted that 92% of them would be severe.) Another report indicated that E1258D had been found in Delta sublineages, but without additional analysis of its potential impact on patient outcomes or transmissibility. [158]

Importantly, E1258D is not lineage specific: 38.7% of the E1258 in the patient data set were from B.1.1.519, 18.1% were P.1 (Gamma), 14.8% B.1.1.7 (Alpha), and 11.9% were B.1.617.2 (Delta) and Delta sublineages (primarily AY.3 and AY.20). As a result, the conventional way of analyzing virulence by studying specific lineages would not pick up on a difference that is due to the presence of E1258D. This demonstrates how sequence modeling can be a crucial compliment to phylogenetic and other lineage-based classifications. E1258D is an example of a mutation that can be adopted, and sometimes can revert, within multiple clades, and different times, and along with different combinations of other mutations. While it is possible that the E1258D mutation’s apparent importance from our deep analysis of the GISAID patient data set may be an artifact of the limited data we analyzed, the strong signal suggests that it may warrant further investigation.

### Genus-Level Coronavirus Classification

The preceding two tasks demonstrate the capabilities of our deep modeling approach to analyze classification and regression problems in the SARS-CoV-2 context. While in these tasks, deep learning can focus on individual sites within the spike protein sequence, our model can be flexibly extended to problems that require deeper sequence analysis. Here, we apply our model, without any changes to structure or hyperparameters, to genus-level classification of coronavirus spike protein sequences. While spike proteins are a ubiquitous feature of coronaviruses, different species in different genera will have spike proteins that bind to different host cell receptors and which have significantly dissimilar structures and sequences. [159], [160] A higher level taxonomic classification task presents, therefore, a qualitative different classification task than one which can succeed by using combinations of single site features.

As a proof of concept, we trained a genus-level Transformer-based neural network classifier for spike protein sequences. We employ the same architecture (from Fig. 1) and the same hyperparameters as in the other studies in this paper, aside from using a softmax function instead of a sigmoid at the output to allow for multiple classes. Because we sought to test our genus-level classifier using SARS-CoV-2 as a validation sample, we removed SARS-CoV-2 and any closely related sequences, i.e., “SARS-like” viruses, recombinant viruses based on SARS-CoV-2, and viruses that were discovered subsequent to the emergence of SARS-CoV-2 specifically to find SARS-CoV-2’s origins (e.g., [161]) from the training data. We then assessed the ability of the trained model to correctly predict that SARS-CoV-2 as *Betacoronavirus*. In a random sample of 100,000 raw, unprocessed sequences, we found that over 10 runs, our model was able to correctly predict 98.4 ± 0.3%.

We also evaluated the model’s ability to classify incomplete and noisy sequences. We found that the model was robust to noise. Of the 36,636 raw sequences in the overall October 1, 2021 data set that contain at least 300 ambiguous amino acid codes, we obtained a correct prediction rate of 90.0 ± 3.8%. The most challenging data set for our model was 14,226 sequence fragments of less than 1000 amino acids in length, where we found a correct rate of 47.7 ± 2.8%. By comparison, BLAST was unable to find hits for 4% of sequences, and in a few cases identified sequences as coming from the wrong genus, and, overall, could correctly identify 95% of short fragment sequences. We do expect that particularly short fragments will challenge our Transformer-based model. Employing a Transformer architecture relies on token and position embedding, since the fragment may be displaced in position in an unpredictable way that cannot be addressed by simple end-padding. However, our results do show that our model can provide robust analysis of sequence patterns even in the presence of a high degree of sequencing noise that results in erroneous or ambiguous amino acid encoding, and it can tolerate shorter lengths of “missing” sequence, such as indels.

The sequence embeddings generated by the genus-level classification model demonstrate how deep sequence modeling can reveal sequence patterns and information at different scales. Fig. 7 shows a t-SNE plot of the embeddings of selected common variants. We can see sequences of certain variants, such as Alpha, Beta, and Delta forming groups. Notably, the patterns that we seem among lineages of SARS-CoV-2 have been found without alignment. Moreover, the only classifier that has been trained is one that classifies at the genus level, without training on any SARS-CoV-2 or closely similar sequences.

**Figure 7.**
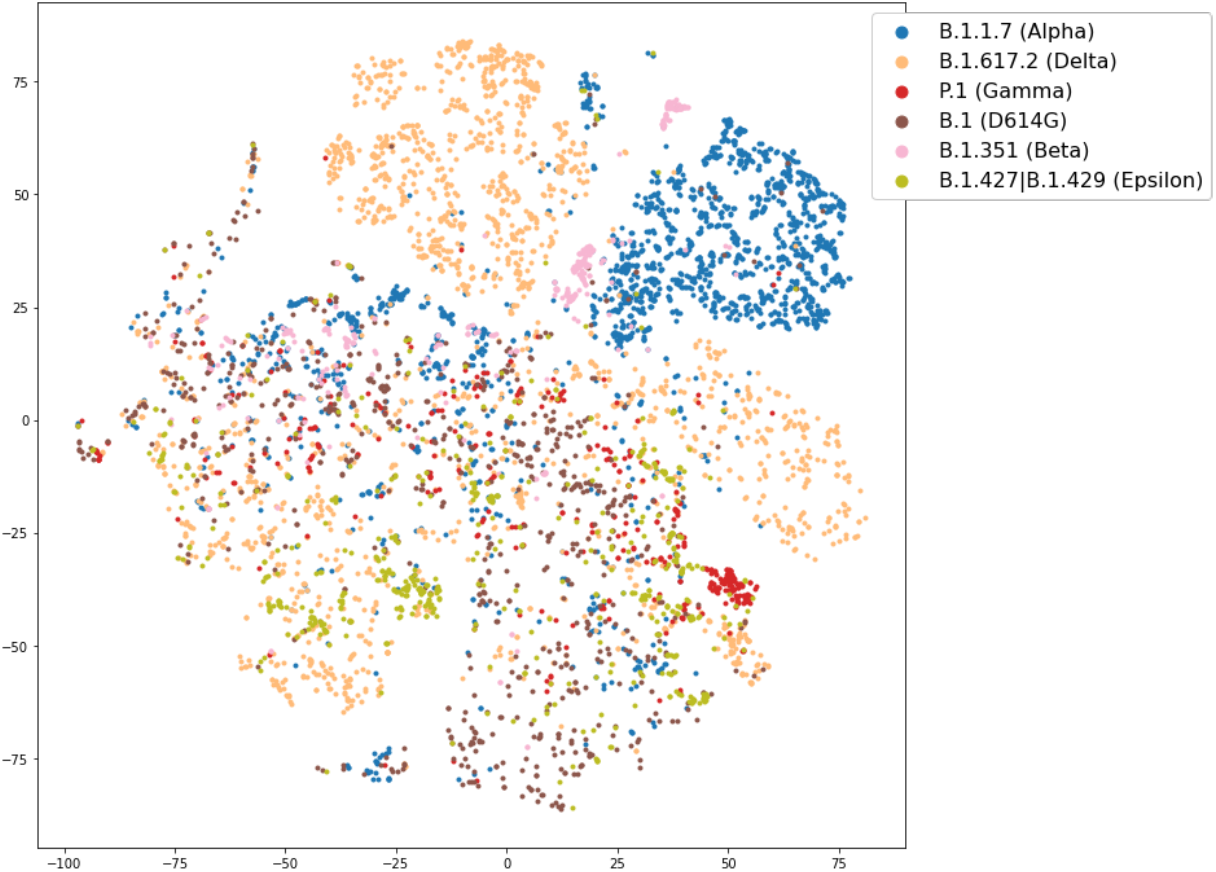
The 2-component t-SNE plot (perplexity of 30) for selected variants in 20,000 raw SARS-CoV-2 spike protein sequences after being classified by the pre-trained genus-level model (with SARS-CoV-2 removed). The selected variants are listed in the legend. (B.1 is ancestral virus with D614G). Despite never seeing SARS-CoV-2, some SARS-CoV-2 sequence embeddings can be clustered by variant, albeit not as well as when classified by time of emergence (Fig. 2).

### Predicting Properties of the Emerging Omicron Variant of Concern

#### Predicting neutralizing antibody reduction (potential immune evasion)

To predict the Omicron variant’s ability to evade the immune responses from vaccination or natural infection, we relied a training data set based on experimental data for the reduction of neutralizing antibody titers due to variants obtained before Omicron. We focus on neutralizing antibody titers, as they have been shown to be a good correlate of immune protection with respect to an infection for vaccines. [162] Table 1 shows the assumptions that we made, which integrate from multiple sources [109]–[113], although we sought to base our training on as few studies as possible to minimize variability. While it may have been better to use data from a single trial, most trials only consider a subset of variants and cannot be used to develop enough training data.

Moreover, our model is fundamentally limited by the very high degree of variability in the source data. Basing a predictive model on neutralizing antibody studies requires contending with noisy measurement methods, substantial inter-individual differences in immune responses reflected in their sera, systematic effects due to using actual virus or pseudovirus (i.e., another virus engineered to express SARS-CoV-2 spike variants), differences arising from whether vaccinated or convalescent sera were used, and differences in antibody reduction due to different vaccines and numbers of doses. [110], [113], [163] For example, in one study, it was found that sera from patients vaccinated with AstraZeneca or Pfizer’s vaccines had on average 8-9 fold lower neutralization, but convalescent sera from patients infected by the ancestral virus (A lineage) had 13.5 fold lower neutralization [164] In sum, our prediction of the reduction of neutralizing antibody activity of Omicron should not be interpreted as an absolute quantitative prediction, rather it will be relative to our starting assumptions in Table 1.

Our initial test set collection consisted of samples annotated as B.1.529 and available on GISAID on December 9, 2021, which, after pre-processing, consists of 295 sequences. We note that while we refer to Omicron and B.1.529 interchangeably, the B.1.529 lineage has more recently supplanted with BA.1, BA.2, and BA.3 designations (described in https://cov-lineages.org/lineage_list.html). We then train our model to perform a regression to the reciprocal of the values in Table 1, as described in the Methods section. Table 5 shows the results for 10 trained models and overall average predictions. There is a high degree of variability between runs, because the predicted neutralizing antibody activity levels are very small: small quantitative differences in predictions due to randomization in the machine learning algorithm result in large variations in the reciprocal prediction of fold-decrease in neutralizing antibodies. Within each run, the model’s predictions also vary between sequences. The relative intersequence variance is consistent within runs, reflecting differences between sequences that are designated as being from the Omicron lineage. Indeed, after these results were obtained, distinct sublineages have been designated within Omicron, i.e., BA.1 and BA.2 sublineages which differ in that BA.2 lacks a deletion at positions 69-70. [165]

**Table 5.**
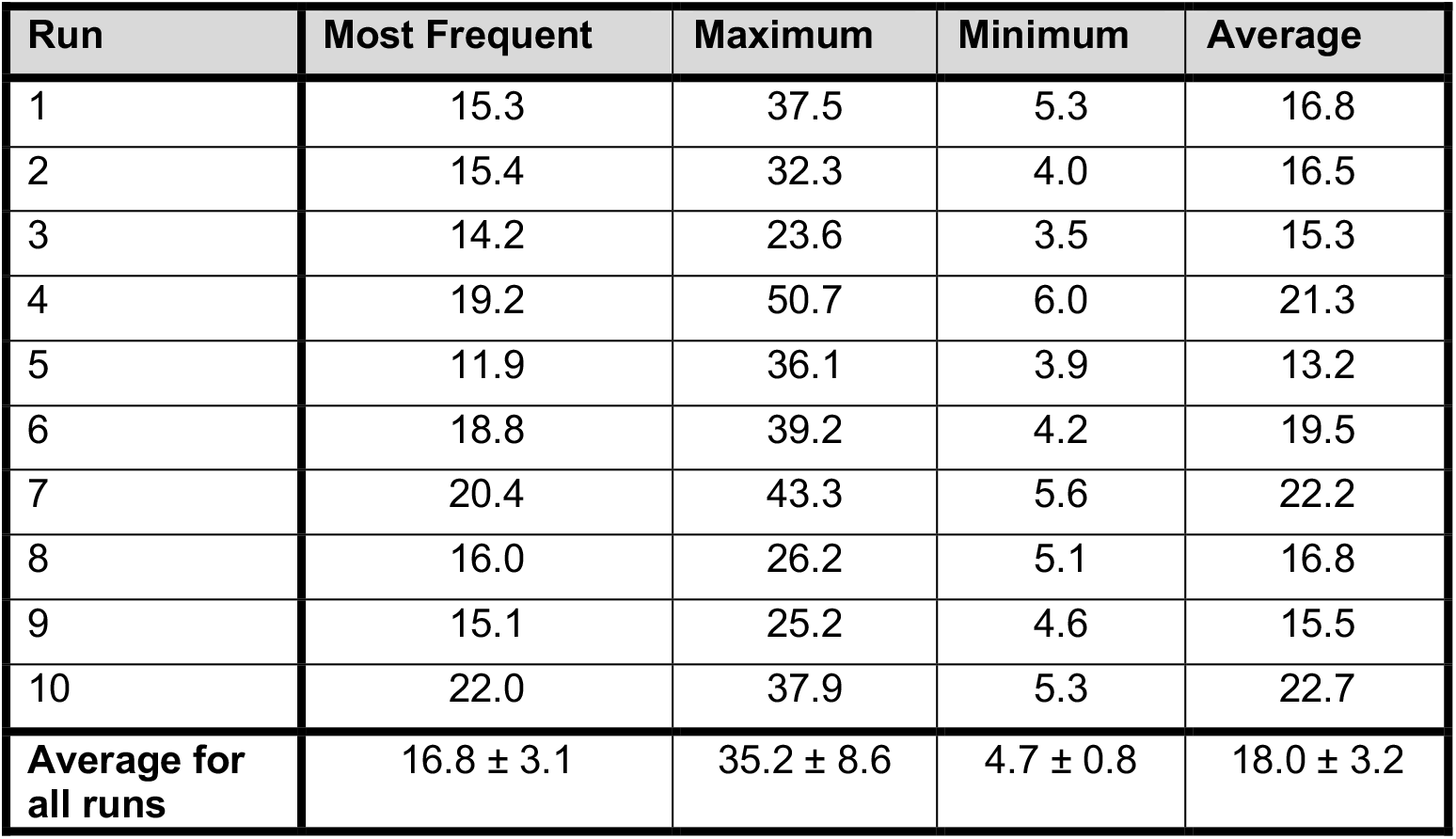
Predicted fold decrease in neutralizing antibody activity for Omicron sequences across multiple runs of the deep learning model. The results show the minimum, maximum, and average predictions across all Omicron sequences, as well as the prediction that was made for the most sequences in the data set. Most runs identified significant decreases in neutralizing antibody activity, even greater than Beta and other variants of concern previously identified as being immune evasive (see Table 1).

| Run | Most Frequent | Maximum | Minimum | Average |
| --- | --- | --- | --- | --- |
| 1 | 15.3 | 37.5 | 5.3 | 16.8 |
| 2 | 15.4 | 32.3 | 4.0 | 16.5 |
| 3 | 14.2 | 23.6 | 3.5 | 15.3 |
| 4 | 19.2 | 50.7 | 6.0 | 21.3 |
| 5 | 11.9 | 36.1 | 3.9 | 13.2 |
| 6 | 18.8 | 39.2 | 4.2 | 19.5 |
| 7 | 20.4 | 43.3 | 5.6 | 22.2 |
| 8 | 16.0 | 26.2 | 5.1 | 16.8 |
| 9 | 15.1 | 25.2 | 4.6 | 15.5 |
| 10 | 22.0 | 37.9 | 5.3 | 22.7 |
| **Average for all runs** | $16.8 \pm 3.1$ | $35.2 \pm 8.6$ | $4.7 \pm 0.8$ | $18.0 \pm 3.2$ |

Notwithstanding the variability in the predictions shown in Table 5, the trends are clear. For certain sequences and runs, the model predicts as high as a 50-fold reduction of neutralizing antibody activity. On average, and for the most frequently observed sequences, Omicron is predicted to have a greater decrease in antibody neutralization than all of the variants previously seen. These results are broadly consistent with both a) predictions that the Omicron spike protein structure can evade antibodies and b) experiments measuring significantly lower neutralization by vaccinated and convalescent sera as compared to previous variants—including Beta, which, as Table 1 indicates, is significantly immune evasive itself. [166]–[171] This demonstrates the ability of the neural network model to generalize, as it predicts a reduction in neutralizing antibody titers of Omicron much greater than any other previously characterized variant considered individually. By contrast, when tested XGBoost an alternative methodology, it consistently overfit to the training data and predicted no more than a 2 or 3-fold reduction for any combination of maximum tree depth, regularization, and other hyperparameters.

We also found that between runs, certain Omicron sequences consistently result in a higher predicted degree of antibody invasion than others. To analyze the mutations that are associated with particularly high levels of immune escape, we determined the difference in the attention scores between the sequence with the largest and smallest predicted fold-decreases in antibody neutralization activity. Fig. 8 shows a plot of the differential attention between maximum and minimum predictions, which shows spikes at positions that are likely to be determinative of the prediction. Certain sites with large differential attention have been characterized as being important for antibody evasion. For example, Fig. 8 shows a spike at position 484, which is where the E484K mutation has been found to lead to substantial antibody escape. [126] Other positions, such as 417, 501, and 681 have also been associated with potential immune-evasive mutations (i.e., N501Y, K417N/T, L452R) [172]. Interestingly, focusing on site 346, we found that a relatively greater reduction in predicted activity was associated with Omicron sequences which include of the R346K mutation. The R346K mutation was found in B.1.621 / Mu variant of concern. [173] Subsequent studies showed reduced neutralizing antibody activity of vaccinated and convalescent sera to the Mu variant. [110] The model’s prediction is consistent with early evidence that an Omicron sublineage with the R346K mutation has substantially less binding affinity to monoclonal antibodies. [169] Other mutations in Omicron sequences that greater neutralizing antibody activity are found at sites 156-158 with an E156G and F157/R158-δ substitution and deletion, which has been linked to a higher level of immune escape in Delta. [174]

**Figure 8.**
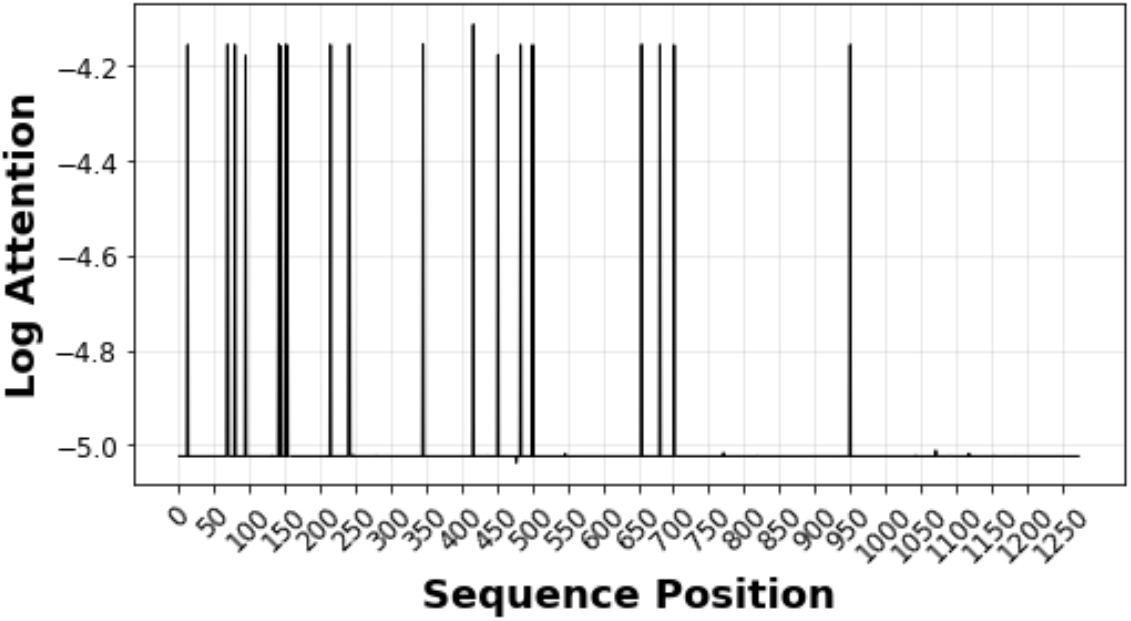
Differential attention plot showing the log (base 10) of the attention difference between the sequence with the maximum and minimum predicted fold-decreases in neutralizing antibody activity (which was the same across all runs shown in Table 5). The spikes show sites that are relevant for predicted increased immune escape, many of which are consistent with sites found to be significant in previous studies.

The model’s results should not be taken as a complete prediction of immune escape, however, because the model does not consider the role of cellular immunity mediated by T cells. Cellular immunity has been shown to likely have a significant role in the immune response to COVID-19 infection and the ability of vaccines to prevent severe disease. [175] For example, patient studies have shown that reduction in neutralizing antibodies, even to the extent of complete elimination of antibodies against the Beta variant, do not necessarily correspond to a drop in real world effectiveness of vaccines with respect to symptomatic and severe disease. [176], [177] Accordingly, vaccines may retain a high degree of protection against severe disease caused by Omicron, even if there were to be a real-world reduction in neutralizing antibody activity to the extent that both the model predicts and recent studies suggest.

#### Predicting Omicron disease severity

As discussed above and shown in Fig. 4C, since Omicron has emerged at a later date in the pandemic than other variants, we expect it to be less severe than if the same variant had emerged earlier in the pandemic. We must therefore control for age and date, as shown Fig. 5D, to compare Omicron’s relative severity to other variants. Fig. 9A shows how the mean predicted disease severity of Omicron sequences would vary over time through the pandemic. Predicted disease severity varies less between sequences than predicted immune evasion; however, there is still some variability as Fig. 9A shows. The maximum standard deviation in the probability of a severe case was found to be less than 0.01 for all time periods, and the largest peak to peak variation is ± 15%. As Fig. 9A illustrates, even the worst-case scenario which would still result in a significantly lower severity than Delta, accounting for the variability of predictions between Delta sequences.

**Figure 9.**
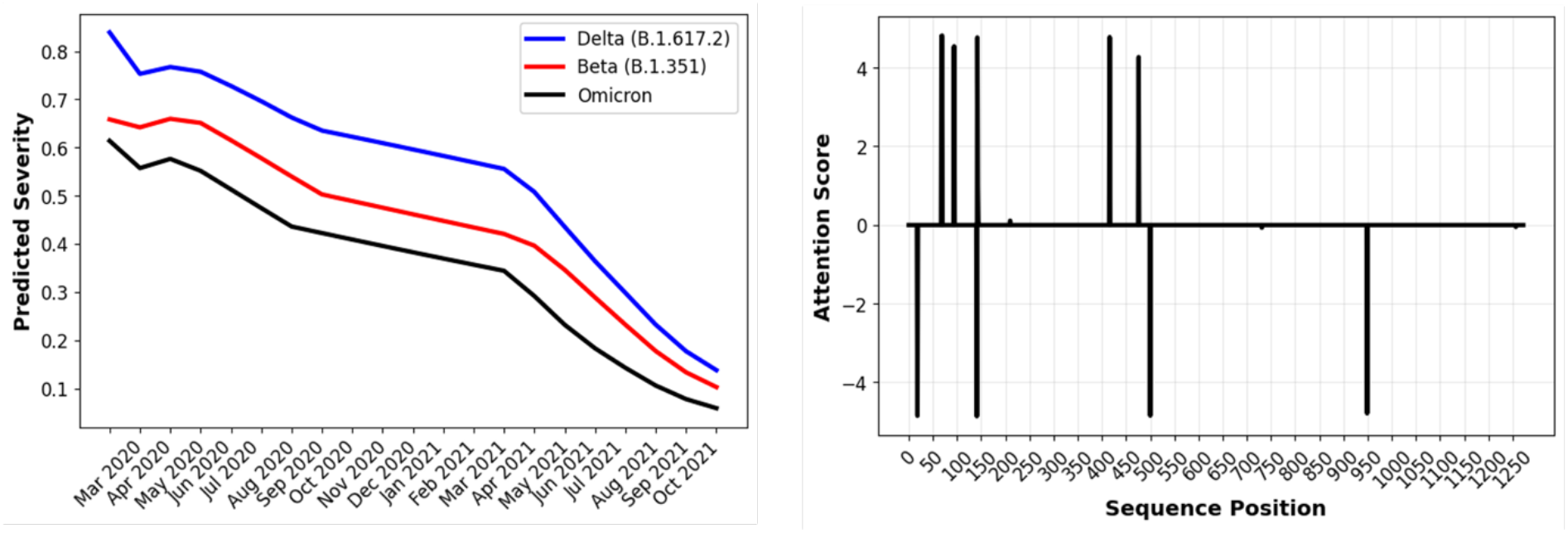
**(A – Left)** Model prediction of the average time-dependent disease severity, assuming a 50-year-old male patient, for samples with known sequences from Beta, Delta, and Omicron lineages. Omicron is predicted to have, on average, a 35-40% reduced risk of severe disease as compared to Delta, and to have reduced risk relative to Beta as well. **(B – Right)** Differential attention plot showing the spike protein sequence positions that have the greatest difference in attention score between most commonly found Delta and Omicron sequences. Sites with a greatest positive attention difference are 69-70 (where Omicron has a deletion also present in Alpha but not Delta), 95 (the most common Omicron sequence type, shown here, lacks Delta’s T95I mutation), 143-144 (Omicron has a deletion), 211 (Omicron has a deletion), 417 (Delta lacks the K417N found in many but not all Delta lineages), and 477 (Omicron lacks S477N). Sites with the greatest negative attention difference are 19 (T19R found in some but not all Delta sequences), 142 (Omicron lacks Delta’s D142G), 501 (N501Y also found in Alpha) and 950 (Omicron lacks N950D).

Fig. 9B shows a differential attention plot, which (similarly to Fig. 8) indicates the spike protein sequence locations that have the greatest difference in attention score between the most prevalent Delta and Omicron sequences. The model found mutations at these sites to be most relevant in predicting that Omicron would likely be less severe than Delta (controlling for patient age, sex, and case date). Further study is required to determine the potential effect of the spike protein sites and mutations identified in Fig. 9B on biological mechanisms that could affect infectivity or cell-cell fusion in ways that would impact the risk of severe outcomes.

The reduced severity of Omicron as compared to Delta is consistent with early laboratory evidence that there is a reduction in lower lung infectivity, deficient cell entry, and a reduction in syncytium formation due to reduced ability of the spike protein to mediate plasma membrane fusion. [178], [179] A decrease in severe disease is also consistent with preliminary clinical and epidemiological evidence that had emerged by the time of this manuscript’s preparation, suggesting that patients had less severe disease in South Africa. [180] Reduced severe disease in South Africa may be due to a particularly high degree of seroprevalence in South Africa as a result of prior infection and vaccination of the vulnerable population. However, early data from the United Kingdom also suggest a 20-25% reduced level of hospitalization of any kind and 40-45% reduction in hospitalizations of greater than one day. [181]

Fig. 9A further shows a reduction in predicted Omicron severity as compared to the Beta variant that had been prevalent in Southern Africa pre-Delta and which also has a high degree of immune evasion. [2] As discussed above, it is difficult to compare clinical and epidemiological evidence of severity for variants earlier in time, because of the effect of vaccination, previous infections, and improved therapies. However, the model’s predictions are consistent with the preliminary data showing that Omicron may have deficient cell entry and less induction of cell-cell fusion as compared to wild type (ancestral genome) as well as Delta. [178] It has also been suggested based on free energy calculations that the Omicron spike protein has deficient ACE2 binding as compared to wild type or Omicron. [182] Even if these and other preliminary results and predictions of lower severity on an individual basis bear out, epidemiological evidence clearly shows that Omicron is highly transmissible, significantly more so than Delta. [183]

Critically, our current model’s predictions of Omicron’s decreased severity depend on accounting for time-dependent factors—in particular, vaccination of the population, improved therapies, and potentially increased opportunities for infection-acquired immunity as well. Fig. 9A shows that had Omicron emerged earlier in the pandemic, absent the protection conferred by vaccination and improved therapeutics, the variant’s apparent severity would have been much greater. Moreover, given the apparently dramatic increased infectivity of Omicron, it may still result in widespread severe disease even at this point in the pandemic.

## CONCLUSIONS

In this paper, we show that a deep sequence model that incorporates a multi-attention Transformer as well as sequence-wide self-attention and embedding layers for multiscale interpretation can successfully perform classification and regression tasks on SARS-CoV-2 spike protein sequence data. Our results demonstrate that our modeling approach can identify attention values that correspond to sequence positions of interest and embeddings that can be visualized to show relationships between sequences. We show that modeling the relationship between disease severity and sequencing variants sets requires accounting for not only patient age but also sample collection date, particularly due to time-dependent increases in vaccination rates. Accounting for these differences by combining demographic variables with sequence input to our model allows us to predict patient severity from GISAID data at nearly 70% accuracy. We can also qualitatively predict that Delta is more severe than Alpha when controlling for time. The same model architecture and parameters used to predict patient severity can be applied to do genus-level classification of coronavirus spike proteins, and embeddings resulting from genus-level classification can be used to visualize lower scale distinctions between SARS-CoV-2 variants.

We have also been able to make predictions about the recently emerged Omicron variant’s immune evasive properties and risk of causing severe disease without using any training data for Omicron. We predict that Omicron will be highly immune evasive, but that it may have substantially less risk of causing severe disease than the Delta variant. Both predictions have so far proven to be consistent with emerging empirical reports. The ability of the model to make validated predictions for Omicron, despite Omicron’s novelty and distance from previously observed SARS-CoV-2 sequences shows that our deep neural network framework is generalizable. We therefore provide the proof of concept for a computational modeling framework that can provide predictive insight on the properties of SARS-CoV-2 variants that may emerge in the future, before empirical data for that variant become available.

Finally, we note that while there is an unprecedented amount of sequencing data available for SARS-CoV-2, the tasks demonstrated here in large part only used thousands of the millions of available samples. This was due to the lack of metadata beyond geographic origin and simple demographic variables for the vast majority of sequences. An important objective of sequencing work should be to collect and curate important information about the sample and to meet minimum information about a sequence standards. [186] However, the fact that we have been able to gain insight to SARS-CoV-2 even with limited data sets shows the potentially for deep modeling approach shown here to be extended to other biological problems.

## Data Availability

All data produced in the present study are available upon reasonable request to the authors, where permitted under GISAID's terms of use. A list of the sequence identifiers (EPI identifiers) for the sequences is available for download from https://github.com/bahrad/Covid. Researchers who register at GISAID website (http://www.gisaid.org) and meet their criteria will gain access to the data. Notebooks used to define models, perform training, and generate results are also available for download from the GitHub site.

## Acknowledgments

We downloaded all SARS-Cov-2 sequences available from and acknowledge the contributions of the Global Initiative on Sharing All Influenza Data (GISAID) EpiCoV database, which has made accessible novel coronavirus sequencing data, including from the NIH Genbank resource. We would also like to acknowledge the authors, originating and submitting laboratories of the sequences from GISAID’s EpiCoV Database on which this research is based, as well as all future SARS-CoV-2 sequence contributors in GISAID’s EpiCoV Database. We particularly acknowledge the researchers from Bostwana, Hong Kong, and South Africa who made the first Omicron genomes available to the community through GISAID. A list of contributors to the data used in this paper will be available at https://github.com/bahrad/Covid. Work reported here was run on hardware supported by Drexel’s University Research Computing Facility.

## Ethics Statement

This study involves only openly available human data, which can be obtained from the GISAID’s EpiCoV database, which is available at http://www.gisaid.org. As this study involves deidentified data made available for public use, it does not require Institutional Review under 45 CFR 46.102.

